# Glycaemic and bodyweight effects of *GIPR* coding variation reflect differences in both surface expression and intrinsic functional impairment

**DOI:** 10.1101/2025.08.15.25333807

**Authors:** Yusman Manchanda, Rofaida Desoki, Eugene J Gardner, John RB Perry, David B Wainscott, Cynthia Stutsman, Claudia Langenberg, Matthew Coghlan, Pallav Bhatnagar, Joseph D. Ho, Michael Yonkunas, Kyle W Sloop, Ken K Ong, Nicholas J Wareham, Alejandra Tomas, Ben Jones

## Abstract

The glucose-dependent insulinotropic polypeptide receptor (GIPR) is the target of several approved and investigational drugs for type 2 diabetes and obesity. Missense coding variation in *GIPR* could confer phenotypic effects through altered constitutive or functional responses to GIP or alter the efficacy of pharmacological agents targeting this receptor. We aimed to provide a deep understanding of the cellular and physiological impacts of individual *GIPR* coding variants and the mechanisms underpinning these effects. By studying a panel of the 30 highest prevalence *GIPR* coding variants in HEK293 cells, INS-1 β-cells and pancreatic islets, we found that many show impaired cAMP responses to physiological GIP stimulation. Population-based association analysis highlighted that these loss-of-function *GIPR* variants decrease BMI but increase glycaemia. In many cases, reduced function was at least partly driven by reduced variant expression at the cell surface due to impaired stability and redirection towards proteasomal degradation pathway. Molecular dynamics simulations suggest distinct variant-induced perturbations in inter- and intra-helical interactions within the transmembrane region which interfere with receptor stability and signalling. This study highlights the mechanisms and consequences of *GIPR* coding variation, which may have implications for the therapeutic targeting of this receptor in metabolic disease.

## 1 Introduction

The glucose-dependent insulinotropic polypeptide receptor (GIPR) is a class B1 G protein-coupled receptor (GPCR) expressed particularly in pancreatic islets, adipose tissue, the brain, and several other tissues (1). Its native peptide hormone ligand, GIP(1-42), is released by intestinal K-cells in response to nutrient ingestion, augmenting glucose-stimulated insulin secretion to regulate post-prandial glycaemia (2). Despite similar anti-hyperglycaemic effects to the related gut hormone glucagon-like peptide-1 (GLP-1) and its pharmaceutical analogues, the GIP system has until recently attracted less attention as a therapeutic target in type 2 diabetes (T2D) and obesity (3). This stems partly from the observation that the insulinotropic effect of GIP is reduced or absent after the onset of T2D (4), but also from the still unclear role for GIPR in bodyweight regulation (5). Counterintuitively, both GIPR agonists and antagonists can promote weight loss in rodents (6, 7), and these paradoxical findings have prompted the development of contrasting anti-obesity strategies in which GLP-1R agonism is combined with either GIPR agonism (e.g. tirzepatide) or antagonism (e.g. maridebart-cafraglutide; MariTide) (8, 9). The observed clinical efficacy of these agents seemingly vindicates these approaches, although how much they depend on GIPR action remains undetermined, particularly as there is a paucity of human data evaluating the effects of sustained pharmacological modulation of the GIP system in isolation.

Understanding of the physiological role of GIPR has been enhanced by genetic evidence accumulated over the past two decades. An early pivotal study demonstrated that *Gipr* knockout mice are resistant to weight gain when fed a high fat diet (10), positioning GIPR antagonism as a potential therapeutic approach for weight loss. However, this effect is not seen when *Gipr* knockout mice are maintained at thermoneutrality (11). *Gipr* knockout mice show mild oral glucose intolerance, reflecting impaired β-cell function (12). Three loss-of-function (LoF) human *GIPR* coding variants – E354Q, E288G and R190Q – were previously reported to reduce BMI (13–16); E354Q is also associated with impaired glucose tolerance and reduced insulin secretion during an oral glucose tolerance test (13, 17), but the effects of the E288G and R190Q variants on glycaemia are less clear. Until recently, phenotypic and pharmacological effects of other *GIPR* coding variants were less well described, especially in comparison with *GLP1R* variants (18–20), but a recent report has provided a broader picture (21). The latter study showed that, with some caveats, LoF *GIPR* mutations typically lead to reductions in both BMI and adiposity but do not influence T2D risk.

Missense coding variation can affect GPCR function via several different mechanisms. The most common cause is reduced delivery of mutated receptors to the plasma membrane (22), and indeed, E288G, R190Q, E354Q, and many of the deleterious variants described in Kizilkaya *et al*. show reduced surface expression (15, 16, 21, 23), as do several *GLP1R* LoF variants (18–20). However, altered function may also result from a change in intrinsic ability to respond to GIP, as has been described for the E288G and R190Q *GIPR* variants (15, 16). In contrast, E354Q shows an “overactivation” phenotype in which accelerated desensitisation is thought to lead to LoF (23). Nuance in how variant GIPRs couple to G protein-dependent and -independent pathways was recently highlighted, as retained coupling to β-arrestin was able to avoid the phenotypic effects of some variants showing loss of cyclic adenosine monophosphate (cAMP) production (21). Importantly, some individual mutations can result in both mislocalisation and intrinsic functional defects, so that disentangling the contribution of each can be challenging. Moreover, deeper molecular and structural understanding of *why* certain *GIPR* variants show reduced surface expression or altered function is lacking. Additionally, except for E354Q (24–26), previous studies of *GIPR* coding variants have used HEK293 and related expression systems that may not fully recapitulate how they behave in their native cellular environment.

In this study we have aimed to address these unanswered questions by performing functional and genetic association analyses of 30 naturally occurring *GIPR* missense variants both in HEK293 and in INS-1 pancreatic β-cells, accompanied by a study of selected variants in primary islets and molecular dynamics simulations to understand the structural basis of the observed effects.

## 2 Results

### 2.1 *GIPR* missense mutations frequently lead to reduced expression at the cell surface

An initial selection of the commonest 30 individual missense *GIPR* variants was made from gnomAD v2.1.1, a dataset which includes whole exome and whole genome sequencing data from large-scale projects involving diverse populations (27). The allele frequency of the rarer variants diverged between different ancestral groups (Figure 1a). Missense variants were distributed throughout the receptor’s extracellular, transmembrane and intracellular domains (Figure 1b).

**Figure 1.**
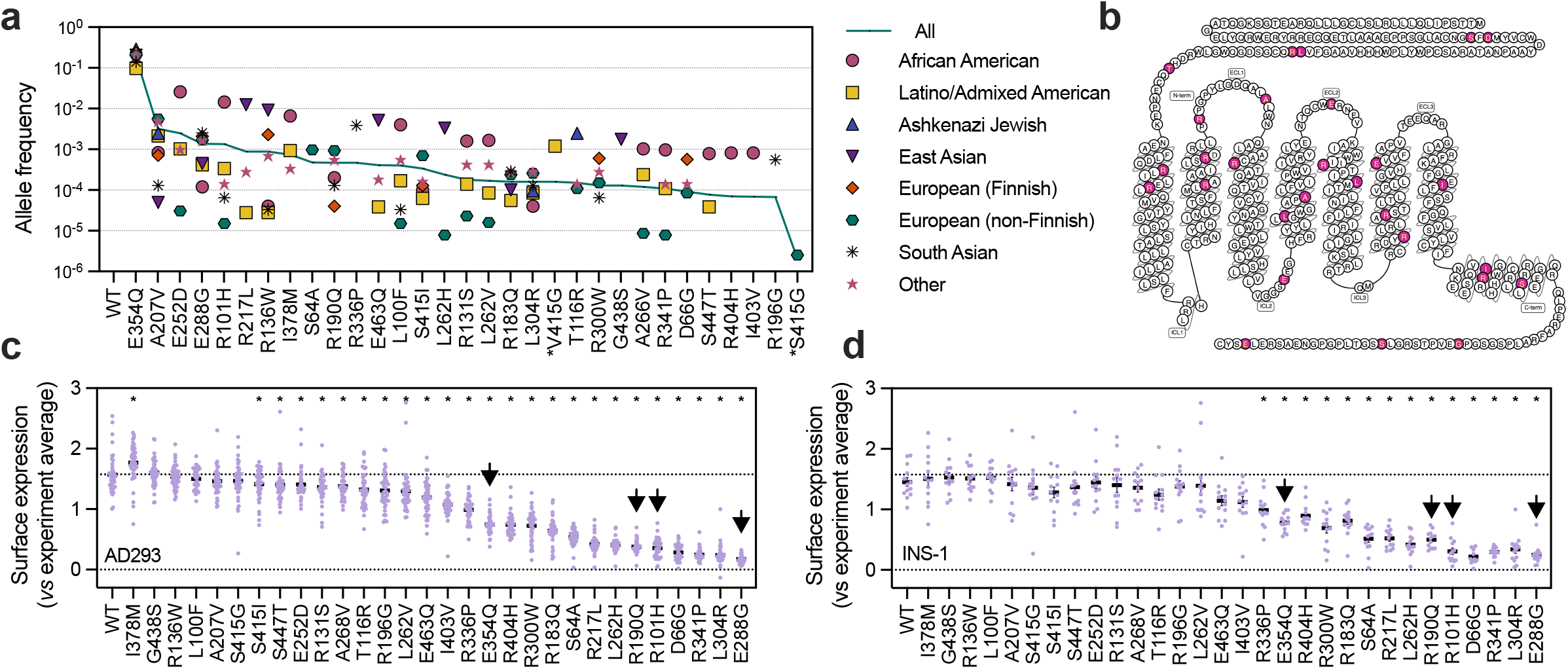
Surface expression levels of GIPR variants in different cellular models. (**a**) Allele frequency of selected *GIPR* variants in different genetic ancestral groups from gnomAD. Note that V415G is higher prevalence than S415G, but as our “wild-type” receptor sequence uses serine at this position, prevalence data for S415G are also shown. (**b**) Positions of selected variants (from gpcrdb.com). (**c**) Mean surface expression of wild-type and variant SNAP-GIPR in AD293 cells, *n*=41, with scaling to the experimental average and comparison by one-way matched ANOVA with Dunnett’s test *versus* wild-type. The dotted line shows the wild-type mean. Arrows highlight E354Q, R190Q, R101H and E288G, which are investigated in more detail below. (**d**) As for (c) but in INS-1 cells, *n*=13. *p<0.05, ****p<0.0001, using indicated statistical test. All data represented as individual data points with mean ± SEM.

To study the impact of these missense mutations on receptor surface expression, we obtained plasmids encoding wild-type and variant GIPR with an N-terminal SNAP tag to allow fluorescent labelling of the receptor. Note that at position 415 major alleles encoding either serine (e.g. variant S415I) and valine (e.g. variant V415G) are mapped (Figure 1a); we used the serine residue as the “wild-type” for functional studies, so henceforth refer to the glycine mutation as “S415G”. We first assessed variant cell surface expression in transiently transfected adherent HEK293 (AD293) cells using a cell impermeant SNAP-tag probe to specifically label GIPR at the plasma membrane (Supplementary Figure 1a). Here, 24/30 variants had a surface expression level that was statistically significantly below that of the wild-type receptor, with 13 of them showing <50% of the wild-type expression level (Figure 1c). Surface expression of the same variants in INS-1 832/3 clonal β-cells devoid of endogenous GIPR after CRISPR/Cas9 deletion (28) (referred to in this work as “INS-1 cells”) was strongly correlated with AD293 results (Figure 1d, Supplementary Figure 1b). Variants previously demonstrated to show reduced surface expression, including E354Q, R190Q and E288G (16, 23), performed as expected in our system. The second and third commonest variants (A207V and E252D) behaved similarly to wild-type GIPR, but the fifth commonest (and second commonest in African American individuals in gnomAD), R101H, showed markedly reduced expression.

We compared our results with those in Kizilkaya *et al*. (21). Of the 13 variants evaluated in both studies, 10 showed good agreement between surface expression measured by SNAP-tag labelling (our study) and B_max_ measured using radioligand binding for untagged GIPR (Kizilkaya *et al*. Supplementary Figure 1c). However, E354Q, A207V and T116R showed divergent results. The discrepancy with E354Q is likely due to experimental variability in the radioligand binding assay as denoted by the reported wide standard error bars (21). Interestingly, additional experiments using SNAP-tagged GIPR in Kizilkaya *et al*. also showed wild-type-like surface expression of the A207V and T116R variants, i.e. similar to our results. We therefore performed radioligand binding assays using a selection of untagged GIPR variants, including A207V, to rule out any interference due to receptor tagging. In our hands, maximal binding for untagged A207V was similar to wild-type GIPR (Supplementary Table 1).

### 2.2 Predicted thermal stability influences variant GIPR surface expression

Reduced surface expression of variant GIPR could result from several mechanisms, for example misfolding during biosynthesis at the endoplasmic reticulum and subsequent degradation, changes to localisation sequences, and accelerated turnover (22). To test for specific deficits in delivery of mature receptor to the cell surface, we labelled “total” SNAP-GIPR using a cell permeable probe, BG-OG (29), alongside standard surface-specific labelling. In AD293 cells, whole cell SNAP-GIPR closely mirrored surface expression levels (Figure 2a), with only small non-significant relative increases in total SNAP detected with the lowest-expressed variants. Moreover, total levels of SNAP-GIPR variants E354Q, E288G, R190Q and R101H, selected for testing due to their relatively higher prevalence and genetically or experimentally confirmed phenotypic effects, were also significantly reduced, as shown by SNAP immunoblot of transfected INS-1 cells (Figure 2b).

**Figure 2.**
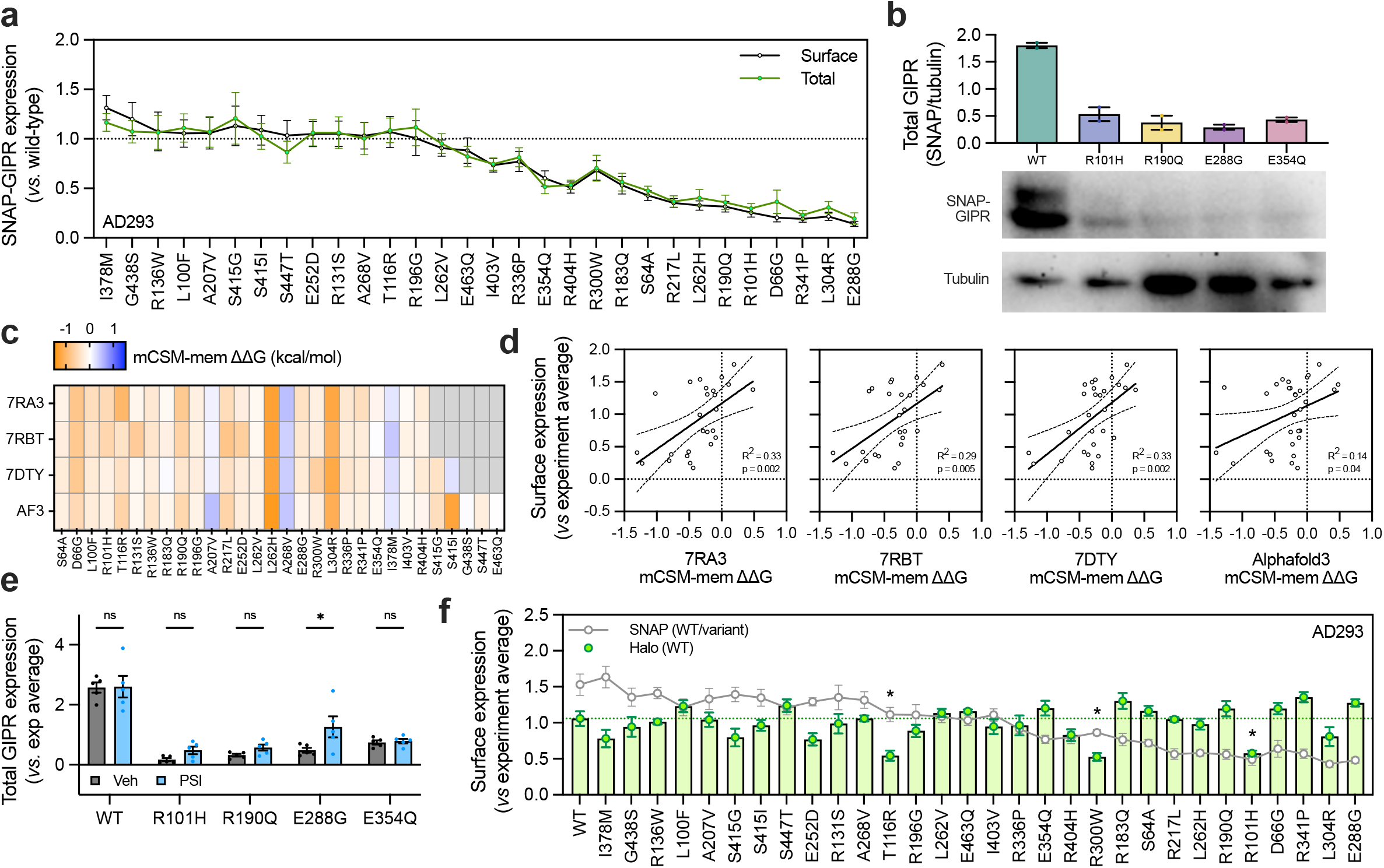
Investigations into mechanisms underpinning altered surface expression of GIPR variants. (**a**) Mean surface and total expression of wild-type and variant SNAP-GIPR in AD293 cells, *n*=8, with normalisation to the wild-type (dotted line) and compared using two-way matched ANOVA with Sidak’s test. See also Supplementary Figure 2a. (**b**) Western blot showing total abundance of wild-type and variant SNAP-GIPR in INS-1 cells, with densitometric quantification of *n*=2. (**c**) Heatmap showing predicted effect on thermostability of each variant as assessed using mCSM-membrane and the indicated experimental (7RA3, 7RBT, 7DTY) and Alphafold (AF3) structures. Decreased stability is predicted when ΔΔG is negative, particularly <-0.5. (**d**) Association between mCSM-membrane-predicted ΔΔG from (c) and measured surface expression in AD293 cells (Figure 1c). The linear regression line with 95% confidence intervals is shown. (**e**) Effect of proteasomal inhibition using PSI (100 µM, 4 hrs) on total SNAP-GIPR levels in INS-1, *n*=5, scaled to experimental average without PSI, and compared using two-way matched ANOVA with Sidak’s test. (**f**) Mean surface expression of wild-type Halo-GIPR and wild-type/variant SNAP-GIPR in AD293 cells, *n*=8, with scaling to the experimental average for each probe, statistical comparison of Halo-GIPR expression in the presence of variant compared to wild-type Halo-GIPR by one-way matched ANOVA with Dunnett’s test. See also Supplementary Figure 2e. All data represented as mean ± SEM with individual data points shown in some cases.

Reduced protein stability is major contributor to pathogenicity of human missense variants (30). Using three experimental GIPR structures (31, 32) and the model predicted by AlphaFold 3 (33), we estimated the impact of each mutation on thermal stability using a tool optimised for transmembrane proteins (34). Three mutations (D66G, L262H and L304R) were predicted to be destabilising (the thermostability effect on the structures as ΔΔG <-0.5) from all four structures, and an additional eight from at least one structure (Figure 2c). In each case, predicted effects on stability showed a significant association with surface expression, with better goodness of fit found with the experimental structures compared to the AlphaFold prediction (Figure 2d). Of note, R190Q and R101H mutations produced ΔΔG <-0.5 with at least one structure, with E288G showing a borderline effect (mean ΔΔG=-0.366 from the experimental structures) and little change for E354Q. As unstable, misfolded nascent proteins in the biosynthetic pathway are frequently redirected from the endoplasmic reticulum towards proteasomal destruction (35), we assessed the effect of 4 hours treatment with a proteasomal inhibitor, PSI (36), on whole cell SNAP levels following transient transfection of SNAP-GIPR variants. Wild-type and E354Q GIPR were unaffected by proteasomal inhibition, but there was a significant increase in E288G levels, with numerically similar trends with R190Q and R101H (Figure 2e). In contrast, inhibition of constitutive GIPR downregulation using the lysosomal acidification inhibitor bafilomycin A1 (37) did not lead to disproportionate “rescue” for GIPR variants with reduced surface expression (Supplementary Figure 2a). These data suggest that reduced plasma membrane levels of selected *GIPR* missense variants result primarily from effects on the biosynthesis or folding of mature GIPR proteins within the cell.

As most *GIPR* genetic variants are encountered in heterozygosity, we also tested for dominant negative effects by co-transfecting wild-type Halo-tagged GIPR with each SNAP-tagged variant and quantifying surface expression of both using spectrally distinct surface-specific probes (Supplementary Figure 2b). Most variants had little effect on wild-type surface expression, but interestingly, three (T116R, R300W and R101H) produced a significant reduction, suggesting dominant negative effects (Figure 2f).

### 2.3 Deciphering the role of *GIPR* variant expression defects on cAMP signalling

As GIPR is canonically coupled to Gα_s_-mediated cAMP signalling, we next measured changes in cAMP by immunoassay in AD293 cells transfected with wild-type or variant GIPR in response to stimulation with 100 pM or 100 nM GIP(1-42), selected to represent physiological (38) and maximal concentrations, respectively. Unsurprisingly, the ten variants with the lowest surface expression showed reduced cAMP responses to both 100 pM and 100 nM GIP (Figure 3a, 3b). However, some of these (e.g. S64A, R217L, L262H) showed at least a partially restored response at the higher ligand concentration. Notably, R136W and R131S showed a reduced response to 100 pM GIP despite close to normal surface expression levels, suggestive of a primary functional deficit. In contrast, E354Q exhibited an increased cAMP response at 100 pM GIP despite reduced expression levels, in keeping with previous knowledge that this variant shows in vitro gain-of-function (GoF) characteristics despite displaying LoF-like physiological impacts (23). E252D and A268V also showed GoF cAMP responses to 100 pM GIP. As the GIPR is thought to have a relatively high rate of constitutive activity (39), we measured basal cAMP from GIPR-expressing cells (i.e. no ligand stimulation); however, there were no differences found between variants and wild-type in AD293 cells (Supplementary Figure 3a). We also measured cAMP responses in INS-1 cells, which showed a similar pattern to that seen in AD293 cells (Supplementary Figure 3b).

**Figure 3.**
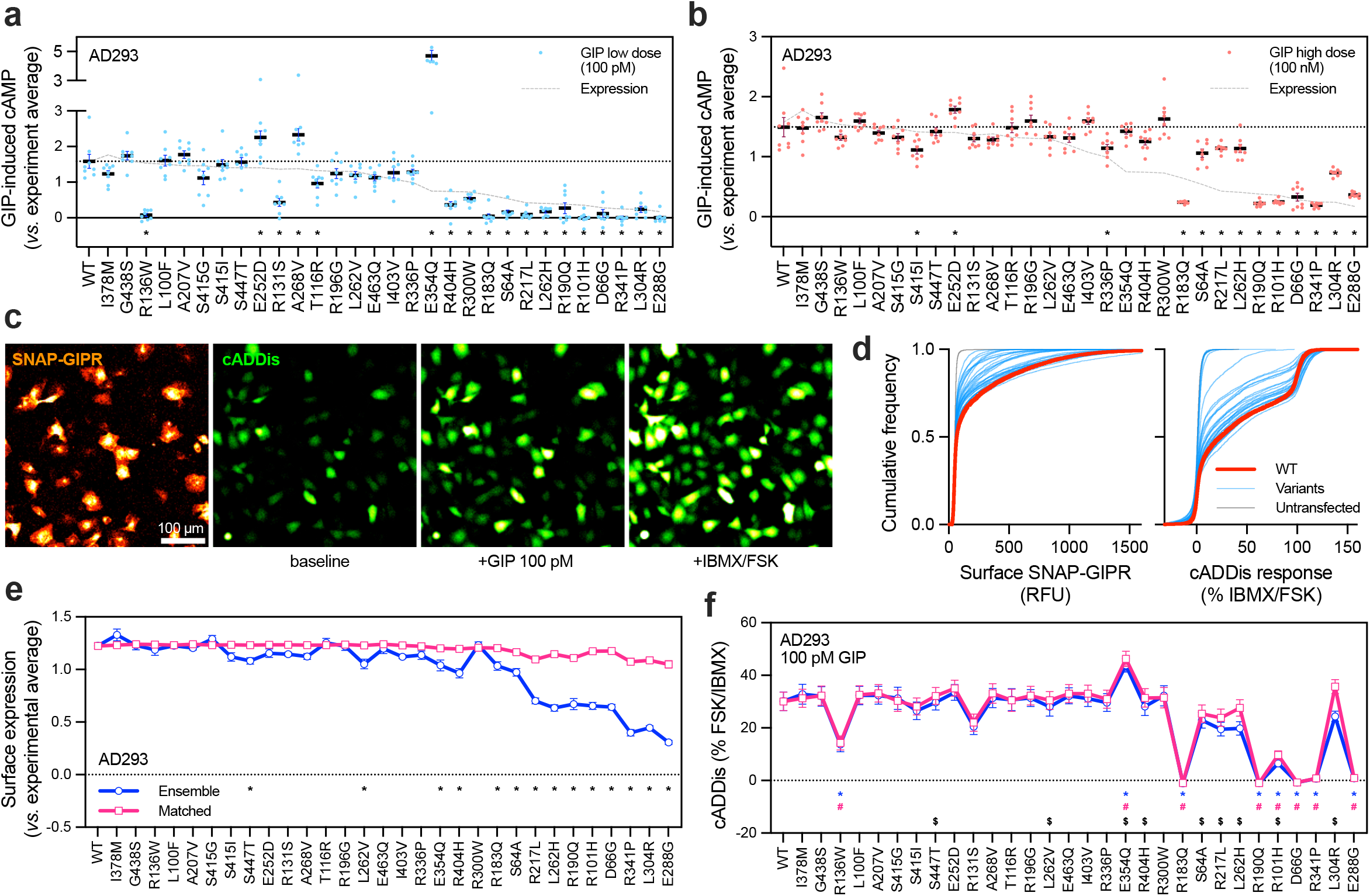
Rescue of cell surface expression does not fully restore cAMP signalling with all variants. (**a**) cAMP responses to 100 pM GIP, measured by HTRF in AD293 cells, *n*=8. Results are scaled to the experimental average and compared by two-way matched ANOVA with Dunnett’s test *versus* wild-type. The dotted line shows the wild-type mean and the dashed line shows the surface expression level from Figure 1c. (**b**) As for (a) but 100 nM GIP. (**c**) Representative image showing cADDis imaging in AD293 cells transfected with SNAP-GIPR and labelled with SNAP-Surface 647. Scale bar = 100 µm. (**d**) Cumulative frequency distributions showing surface GIPR labelling and cADDis responses from a single experiment. (**e**) Surface GIPR expression from ensemble (unselected) and expression-matched analysis of cADDis experiment, *n*=8, with scaling to the experimental average and comparison by two-way matched ANOVA with Dunnett’s test *versus* wild-type. See also Supplementary Figure 3e. (**f**) cADDis response to 100 pM GIP from same experiments as (e), with two-way matched ANOVA with Dunnett’s test *versus* wild-type, and Sidak’s test comparing ensemble and expression-matched data. See also Supplementary Figure 3f. *p<0.05 (variant *versus* wild-type from ensemble cells), #p<0.05 (variant *versus* wild-type from expression-matched cells), $p<0.05 (ensemble *versus* expression-matched cells). All data represented as mean ± SEM with individual data points shown in some cases.

Whilst these data demonstrate a clear overall positive association between GIPR variant surface expression and cAMP response (Supplementary Figure 3c), they provide an incomplete description of the deleterious impact of certain variants, as impaired surface expression could disguise an additional functional deficit. Elucidating the relative contribution of each is potentially important as mutations that primarily affect trafficking to the plasma membrane could be amenable to rescue e.g. by pharmacological chaperones (40). To resolve this question, we used the cell-to-cell heterogeneity in plasmid uptake during transient transfection to facilitate biosensor recordings of cAMP responses from individual cells with different levels of surface GIPR. This high content imaging assay involves fluorescence imaging of thousands of cells in parallel after co-transduction of SNAP-tagged GIPR and the live cell cAMP sensor cADDis (41) (Figure 3c, 3d). cAMP responses to 100 pM GIP from “ensemble” (non-selected) AD293 cells measured in this assay agreed with the biochemical cAMP measurements, although the relationship was non-linear due to the greater sensitivity of the Epac2-based biosensor (Supplementary Figure 3d). We used a weighting procedure based on grouping variant-expressing cells into deciles matched to the surface expression of the wild-type, allowing cAMP responses to be ascertained at equalised surface expression levels from different receptor variants (Figure 3e, Supplementary Figure 3e). Notably, all variants with reduced ensemble cAMP responses compared to wild-type still showed LoF after matching for surface expression levels, indicating that they all have intrinsic defects in cAMP generation (Figure 3f, Supplementary Figure 3f). However, for some of these variants there was only a partial restoration of response, e.g. S64A, R217L, L262H, R101H and L304R. We interpret this to indicate that, for these variants, cAMP LoF reflects both reduced surface expression and intrinsically impaired GIP-responsiveness. In contrast, other variants such as R183Q, R190Q, D66G and E288G showed no rescue of function even at wild-type-like expression density. The same analysis performed using a saturating concentration of GIP (100 nM) GIP showed that expression matching only very partially rescued function for the latter four variants, highlighting their profound functional defects (Supplementary Figure 3g-j).

### 2.4 Limited effects of missense variants on GIPR internalisation

Preserved β-arrestin-mediated GIPR internalisation has been suggested to influence *GIPR* missense variant phenotypic effects by driving endosomal signalling (21). We used a reversible SNAP-tag labelling technique (Figure 4a) to quantify both constitutive and GIP-induced internalisation of wild-type SNAP-GIPR in AD293 cells and INS-1 β-cells. Of note, 40% of wild-type GIPR was constitutively internalised (i.e. without agonist) over 60 minutes in AD293 cells, increasing to approximately 60% in the presence of a maximal concentration of GIP (Supplementary Figure 4a). Thus, an additional 20% of surface-labelled GIPR underwent internalisation when agonist was applied at a saturating concentration, with an EC_50_ almost 300 times higher than for cAMP production in this cell model (Supplementary Figure 4b), in keeping with previous reports highlighting the relatively limited capacity for GIP-induced GIPR internalisation (42, 43). It should be noted however that, despite this relatively small net increase in internalisation, the majority of receptor internalised at this saturating concentration will be bound to GIP.

**Figure 4.**
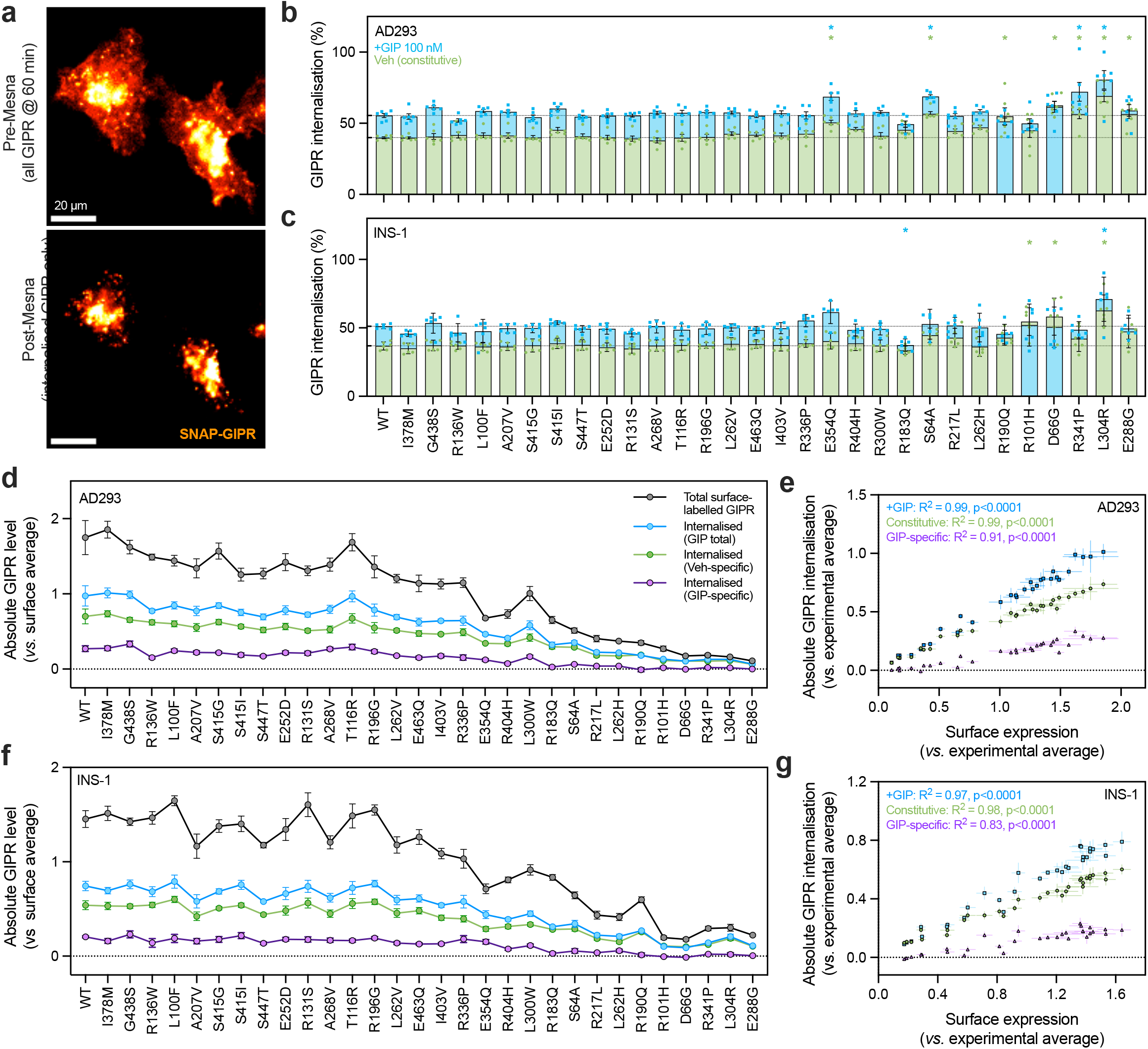
Constitutive and GIP-induced GIPR internalisation effects. (**a**) AD293 cells expressing wild-type SNAP-GIPR, pre-labelled with BG-SS-649, and treated for 60 minutes with 100 nM GIP; imaging performed before and after Mesna treatment. Scale bar = 20 µm. (**b**) Percentage of pre-labelled SNAP-GIPR internalised over 60 minutes with vehicle or 100 nM GIP treatment in AD293 cells, *n*=6, with comparison of variants *versus* wild-type by two-way matched ANOVA with Dunnett’s test. Dotted lines represent mean wild-type constitutive and GIP-stimulated responses. (**c**) As for (b) but in INS-1 cells, *n*=7. (**d**) Quantification of absolute amount of total and internalised surface-labelled GIPR in AD293 from the experiments shown in Figure 4b, highlighting the relative contributions of constitutive and GIP-specific internalisation. Results have been scaled to the experimental average for total surface GIPR expression. (**e**) Relationship between AD293 surface GIPR expression from Figure 1a and absolute amount of GIP-induced GIPR internalisation amount from Figure 4d. (**f**) As for (d) but using INS-1 data from Figure 4c. (**g**) As for (e) but using INS-1 data from Figure 1b and Figure 4f. *p<0.05 by indicated statistical test. All data represented as mean ± SEM with individual data points shown in some cases.

Most *GIPR* missense variants showed wild-type-like internalisation in the presence of 100 nM GIP, with similar results with AD293 and INS-1 cell models (Figure 4b, 4c, Supplementary Figure 4c). Some moderate increases in internalisation were recorded with selected low-expression variants, such as E354Q, S64A, R341P, L304R and E288G, which was driven by apparent increases in constitutive turnover. However, it should be emphasised that earlier experiments with bafilomycin A1 (Supplementary Figure 2a) argue against accelerated constitutive downregulation being a primary driver of reduced surface expression with these *GIPR* variants. When the GIP-induced increase in total internalisation was analysed separately, several variants including R190Q, R101H, E288G, D66G and R183Q showed statistically significant reductions in internalisation in either/both AD293 and INS-1 cells (Supplementary Figure 4d). This provides further support for impaired intrinsic function of these variants, as suggested from the cAMP results in Figure 3f.

Whilst percentage of internalisation is helpful to understand functional deficits, the absolute amount of internalised GIPR may also be informative. In our analysis, it was both clear and unsurprising that the amount of GIPR internalisation (constitutive and GIP-mediated) was strongly dependent on variant-specific surface expression levels (Figure 4d-g). As capacity for endosomal signalling is, by definition, influenced by the total bulk of internalised agonist/receptor complexes, differences in surface expression are likely to play a major role in controlling the relative ability of *GIPR* variants to generate signals from intracellular compartments.

### 2.5 Molecular dynamics simulations provide insights into structural basis for variant effects

Having demonstrated that several GIPR variants show both impaired surface expression (perhaps due to thermodynamic instability) and also compromised ability to respond to GIP, we aimed to understand the possible structural basis for these effects. Microsecond scale molecular dynamics (MD) simulations based on a wild-type GIP-bound structure and four prioritised mutations (R190Q, E354Q, E288G, and R101H) were used to assess a potential structural impact at the molecular level.

While none of these variants make direct hydrogen bonding interactions with the GIP peptide, several residue-residue interaction frequencies were chosen as observables to monitor the interhelical stability of the transmembrane peptide binding domain over simulation time. R190Q disrupts both local and distant inter-helical interactions within the GIP binding site. The local intra-helical interaction between wild-type R190 and D191 is completely lost in the presence of this variant (Figure 5a). This change also disrupts the TM1:Y141 – TM2:D191 interhelical interaction observed in the wild-type receptor. Additionally, the interhelical interaction between TM4:Y231 and TM6:E354 is moderately disrupted, suggesting long-range effects of this mutation. These long-range disruptions suggest that not only the size of the residue in position 190 is important but also the change and are consistent with experimentally measured reductions in surface expression and cAMP production. E288G produces intra and inter-helical interactions like that of the wild-type APO receptor (Figure 5b). The most significant change in residue-residue interactions comes from the central R190-D191 interaction on TM2. Microsecond simulations show a movement of the GIP peptide across the receptor that reduces the Pi-cation interactions of GIP:F6 and GIP:I7, as well as engage in a new hydrogen bond between the backbone carbonyl of E288G and GIP:S8 sidechain hydroxyl (Supplementary Figure 5b). This result is consistent with observed variant expression defects on cAMP signalling, noting E288G’s only partly rescued function even at 100 nM GIP. E354Q shows moderate disruption of protein-protein interactions (Figure 5c). The TM4:Y231 – TM6:E354 interaction is key to stabilizing the GIPR bundle in the presence of GIP, shown from the high frequency hydrogen bonding and hydrophobic interactions between residues in the wild-type simulations. Introducing the E354Q mutation reduces but does not destroy this inter-helical interaction. Long range effects are also minimal with a slight enhancement in the TM1:Q138 – TM2:D191 interaction. R101, located in the ECD, shows minor disruption of long-range protein-protein interactions at TM4-TM6 in the R101H variant form (Supplemental Figure 5c). While R101H does not disrupt the local orthosteric interactions typically observed in the presence of GIP, there is a small consistent long-range effect. All the variant simulation data suggest the TM6 E354 – TM4:Y231 inter-helical bundle interaction is sensitive to changes in receptor sequence. Since E354 is located on TM6, directly implicated in signal transmission through the TM6 kink, even long distance (both sequentially and spatially) could influence G protein engagement.

**Figure 5.**
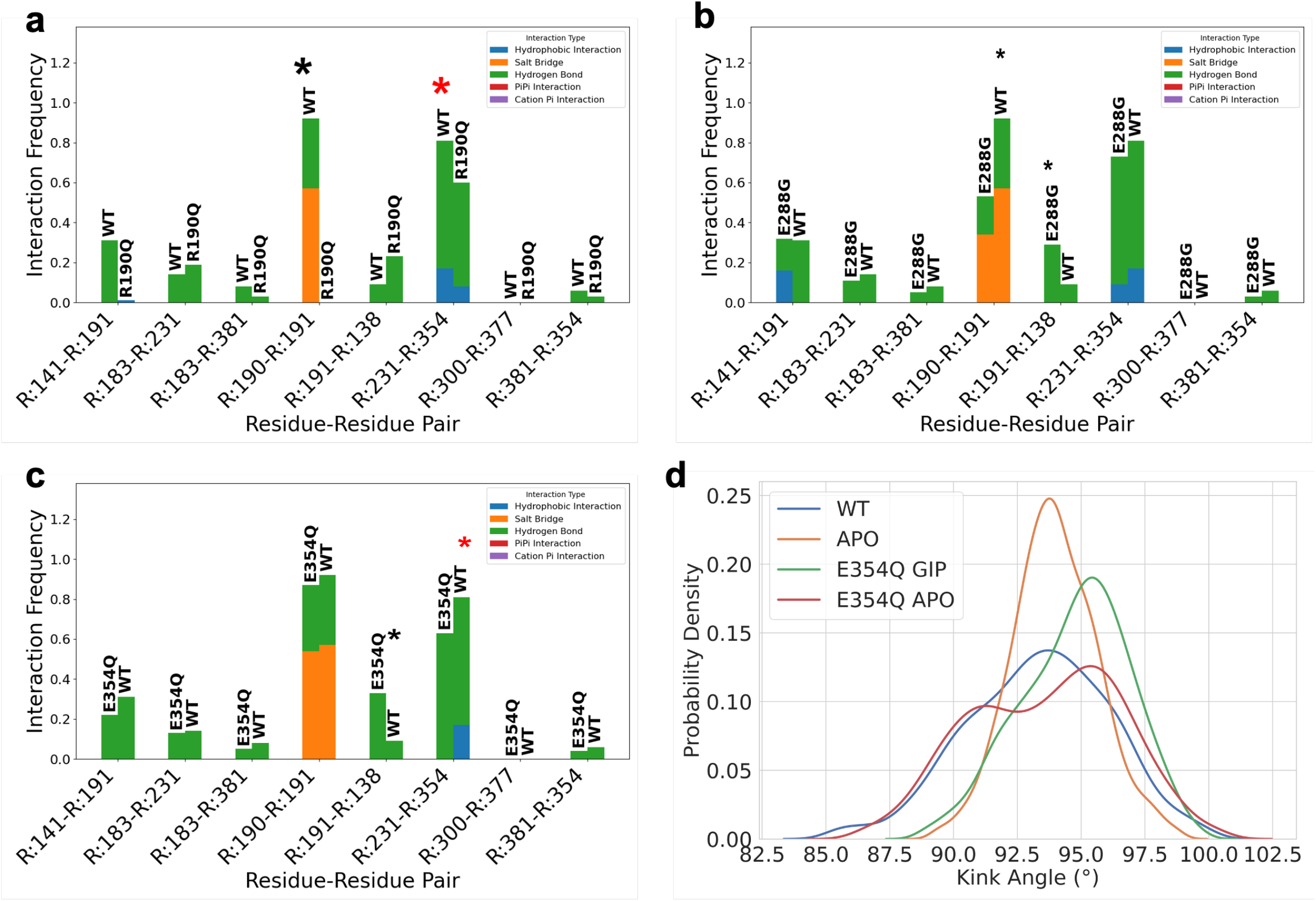
Protein-protein interactions from molecular dynamics simulations. (**a**) Mutation R190Q disrupts both local and distant inter-helical interactions within the GIP binding site. The local intra-helical interaction between R190 and D191 is completely lost in the presence of the Q190 mutant. This change also disrupts the TM1:Y141 – TM2:D191 interhelical interaction observed in the wildtype receptor. The interhelical interaction between TM4:Y231 and TM6:E354 is also moderately disrupted suggesting long range effects of this mutation. (**b**) Mutation E288G produces intra and inter-hekical interactions like that of the wildtype APO receptor (see Supplementary Figure 5a). The most significant change in residue-residue interactions comes from the central R190-D191 interaction on TM2. Microsecond simulations show a movement of the GIP peptide across the receptor to reduce the Pi-cation interactions of GIP:F6 and GIP:I7; as well as engage in a new hydrogen bond between the backbone carbonyl of E288G and GIP:S8 sidechain hydroxyl (see Supplementary Figure 5a). (**c**) Mutation E354Q show moderate disruption of wildtype protein-protein interactions. The TM4:Y231 – TM6:E354 interaction is key to stabilizing the GIPR bundle in the presence of GIP shown from the high frequency hydrogen bonding and hydrophobic interactions between residues in the wildtype simulations. Introducing the E354Q mutation reduces but does not destroy this inter-helical interaction. Long range effects are also minimal with a slight enhancement in the TM1:Q138 – TM2:D191 interaction. (**d**) TM6 kink-angle defined by the vectors from residues 343 – 348 and 348 – 353. The shift in probability from 93.7^°^ (more kinked) to 95.6^°^ (less kinked) for the wild-type and E354Q variant respectively indicate a decrease in probability of finding the receptor in the active state in the presence of native peptide.

To understand the role that the TM6:E354 – TM4:Y231 inter-helical interaction, the E354Q variant, and modulators thereof play in protein stability and dynamics, the angle defined by the vectors from residues 343 – 348 and 348 – 353 were used to represent the TM6 “kink”. A probability distribution was calculated over simulation time as a function of this angle for both wild-type and the E354Q variant (Figure 5d). The shift in peak probability from 93.7^°^ (active reference) to 95.6^°^(less kinked, less active) for the wild-type and E354Q variant respectively suggest lower probability of finding the receptor in the active state in the presence of native peptide. Interestingly, the APO E354Q variant exhibits a modified probability distribution compared to the wild-type APO receptor resembling that of a GIP bound wild-type receptor. The bi-modal distribution is characteristic of a more dynamic system, and points towards a potential increase in constitutive activity, as hinted at by moderately increased basal turnover of this variant in our internalisation assays (Figure 4).

### 2.6 Assessment of *GIPR* variant function in pancreatic islets

To build on our functional screening results from AD293 and INS-1 cells, we examined the effects of selected variants in a primary pancreatic islet environment. Focussing on higher prevalence variants with altered function in cellular models, we confirmed the expected surface expression patterns of E354Q, R190Q, R101H and E288G using adenoviral SNAP-GIPR expression in mouse islets, with marked reductions seen particularly with the latter three variants (Figure 6a, 6b).

**Figure 6.**
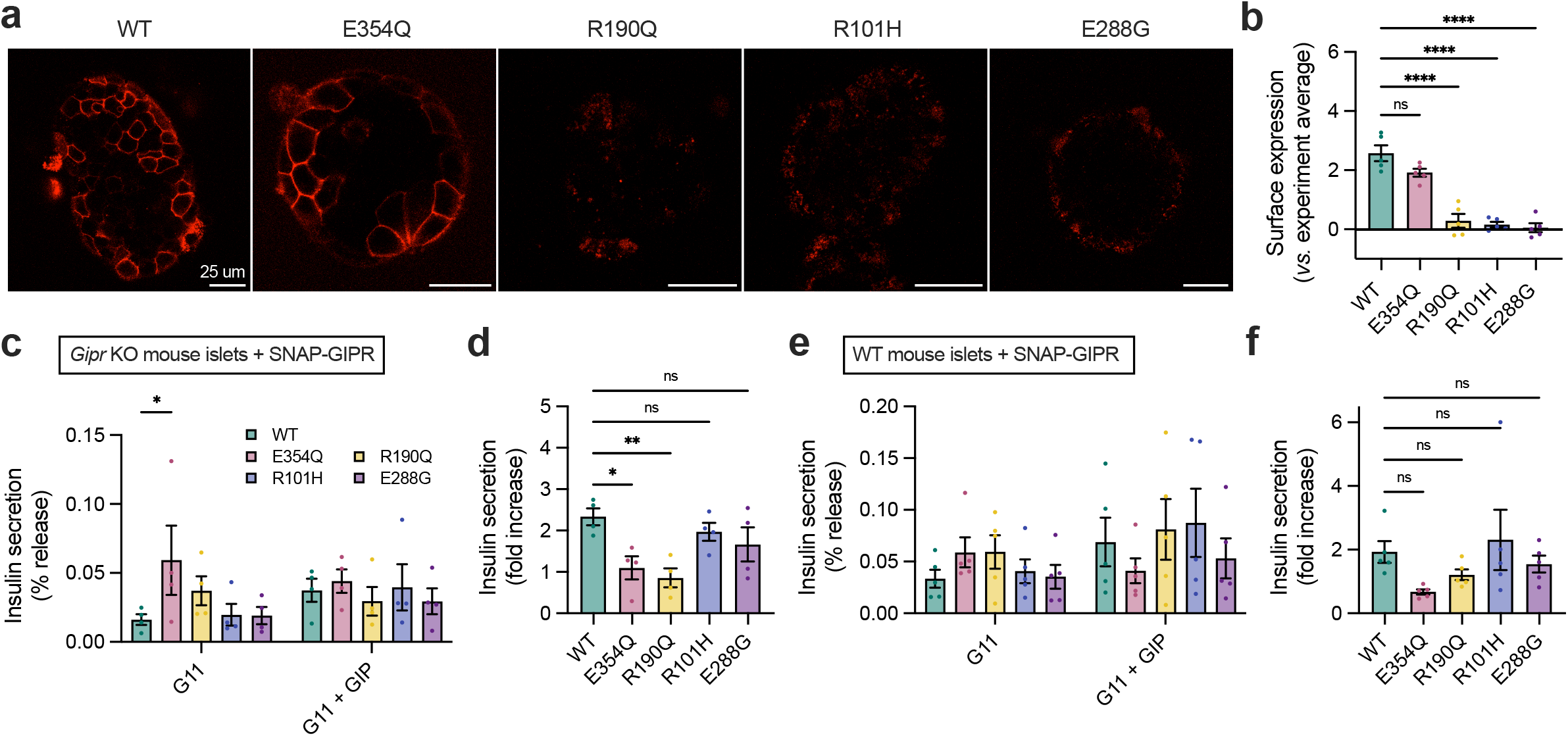
Evaluating variant effects in pancreatic islets. (**a**) Representative confocal images showing surface labelling of wild-type and variant SNAP-GIPR in mouse pancreatic islets after adenoviral transduction. (**b**) Quantification of surface SNAP-GIPR from *n*=5 repeats and comparison by one-way matched ANOVA with Dunnett’s test *versus* wild-type. (**c**) Release of insulin from *Gipr*^-/-^ mouse islets transduced with GIPR adenoviruses at 11 mM glucose (G11), with and without 100 nM GIP, 30-minute stimulation, *n*=4. Wild-type and variant responses compared by two-way matched ANOVA with Dunnett’s test. (**d**) Data from (c) analysed as GIP-induced fold-change, with comparison using one-way matched ANOVA with Dunnett’s test. (**e**) As for (c) but in wild-type mouse islets after adenoviral transduction, *n*=5. (**f**) As for (d) but analysing insulin data data from (e). *p<0.05, ****p<0.0001, using indicated statistical test. All data represented as mean ± SEM with individual data points.

GIPR is expressed in both pancreatic β- and α-cells, where it regulates release of insulin and glucagon, respectively (44). We performed hormone release assays in islets from *Gipr* knockout mice transduced with wild-type or variant GIPR, to examine secretory effects in the absence of the endogenous receptor. We found that E354Q expression led to a significant increase in constitutive insulin secretion at 11 mM glucose (Figure 6c), compatible with a higher constitutive activity level of this variant as hinted at from the internalisation assays shown in Figure 4 and the MD results in Figure 5d. However, E354Q showed a blunted insulinotropic response to GIP stimulation compared to wild-type GIPR (Figure 6d), in keeping with the reduced C-peptide response to oral glucose reported in people carrying this variant (17). R190Q, R101H and E288G showed apparent reduced GIP-induced insulin secretion, although this was statistically significant only for R190Q. Each variant also showed apparent but non-statistically significant reduction in glucagon release at 0.5 mM glucose compared to wild-type (Supplementary Figure 6a, 6b). We also assessed responses in islets from wild-type mice, in which the mouse GIPR is expressed endogenously (Figure 6e, 6f). Although not statistically significant, the same trends were observed, with a tendency towards reduced responses to GIP stimulation particularly with E354Q and R190Q. Binding of the fluorescent GIP analogue GIP-TMR (43) was also tested in wild-type islets transduced with SNAP-GIPR adenovirus, showing high levels of binding to wild-type and the E354Q variant but very low levels with the other variants which were numerically reduced compared to even non-transduced control, hinting at possible dominant negative effects when over-expressed (Supplementary Figure 6c).

### 2.7 Population-based validation of *GIPR* variant effects

To assess the phenotypic impact of the 30 *GIPR* missense variants we turned to UK Biobank (UKBB), in which 28 of our 30 functionally characterised variants were represented. We performed rare variant burden testing using STAAR (45) to identify potential associations with BMI, glycated hemoglobin levels (HbA1c, in individuals without T2D), and T2D of distinct *GIPR* variant groups classified according to our in vitro data – specifically the 100 pM GIP cAMP responses and levels of surface expression in AD293 cells. We assigned variants to reduced, neutral and increased response groups, with “reduced” defined as a statistically significant reduction of at least 50% below that of the wild-type receptor and “increased” as a significant increase above wild-type. BMI was included as a covariate to determine associations with T2D and HbA1c independently of BMI. The common variant E354Q was excluded from burden tests due to its high frequency. All other individual variants had a MAF < 0.2% in the full multiethnic UKBB sample. Results on Europeans-only and full multiethnic UKBB samples were consistent, and statistical power was enhanced in the latter by its larger sample size and inclusion of rare variants that are more prevalent in non-Europeans. Results were also consistent in generalised linear regression sensitivity models. The following results are from STAAR models in the full multiethnic UKBB sample. Results from all burden testing analysis models are presented in Supplementary Table 2.

As summarised in Figure 7a, rare *GIPR* variants that conferred reduced cAMP signalling (14 variants, up to 3,867 carriers) were collectively associated with lower BMI (P=8.4×10^−10^) but higher BMI-adjusted HbA1c (P=3.9×10^−8^), with a nominal association with higher BMI-adjusted T2D risk (OR=1.21, P=4.1×10^−3^). By contrast rare *GIPR* variants with neutral effects on cAMP signalling (11 variants, up to 7,047 carriers) showed only nominal collective association with lower BMI (P=2.8×10^−2^) and no association with HbA1c (P=7.4×10^−1^) (Figure 7a). No collective association was found with the two rare variants that conferred increased cAMP responses (E252D, A268V), although a directionally consistent trend towards reduced HbA1c and T2D risk was seen. The associations of *GIPR* in vitro LoF with increased HbA1c and T2D risk were consistent with or without adjustment for BMI (Supplementary Figure 7a). Similar findings were observed when categorising variants according to surface expression, with lower surface expression variants (11 variants, up to 3,656 carriers) associated with reduced BMI (P=7.0×10^−11^), and increased risks of BMI-adjusted HbA1c (P=3.6×10^−7^) and BMI-adjusted T2D (P=3.5×10^−3^) (Supplementary Table 2).

**Figure 7.**
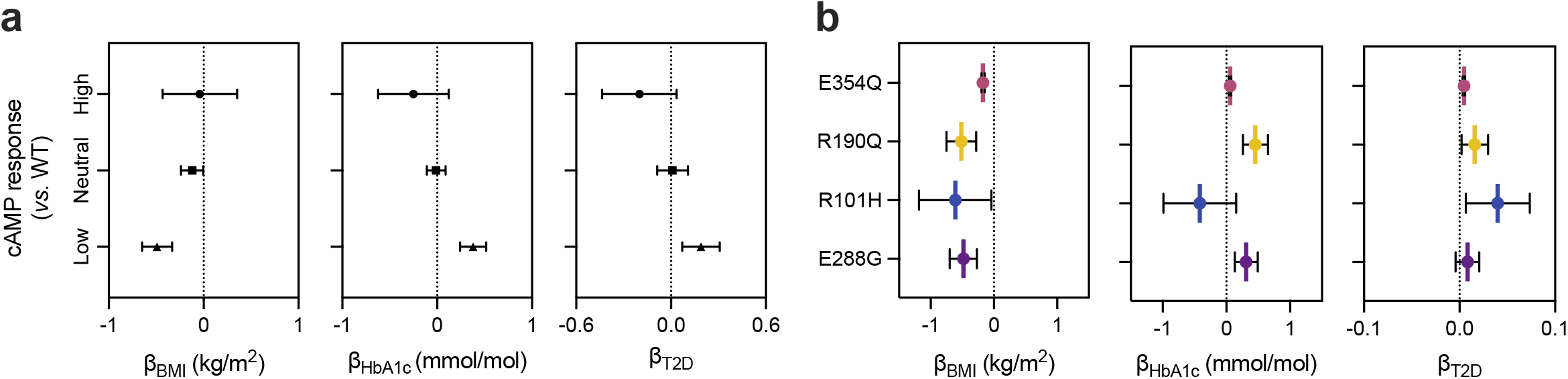
Contrasting effects of GIPR missense variants on BMI and glycaemic status. (**a**) Results of burden test analysis, showing effects of *GIPR* variants grouped by cAMP response category on BMI, HbA1c (BMI-adjusted) and T2D risk (BMI-adjusted). (**b**) Results of selected single variant association analyses, showing effects of E354Q, R190Q, R101H and E288G on BMI, HbA1c (BMI-adjusted) and T2D risk (BMI-adjusted). Data are represented as mean ± 95% confidence intervals.

To assess individual variant impacts on the same outcomes, we performed single-variant association testing in the full multiethnic UKBB WES sample using BOLT-LMM v2.4.1 (46), with BMI included as a covariate for T2D and HbA1c associations, as above. Results from the four commonest LoF variants (E354Q, R190Q, R101H, E288G) are shown in Figure 7b, with full data in Supplementary Table 3. In keeping with previous reports (13), the common E354Q variant (MAF=19%, up to 178,179 carriers), showed statistically robust but relatively modest effects on decreased BMI (P=3×10^−49^) and increased BMI-adjusted HbA1c (P=3.2×10^−9^) and T2D risk (P=8×10^−12^), directionally-consistent with the above LoF variant groups. E288G (MAF=0.18%, up to 1,715 carriers) and R190Q (MAF=0.15%, up to 1,373 carriers) also showed significant effects on decreased BMI (P=6×10^−6^ and P=1.6×10^−5^, respectively) and increased BMI-adjusted HbA1c (P=6.9×10^−4^ and P=9.4×10^−6^, respectively), but with larger effects than E354Q, indicative of more dramatic impacts on receptor function. Even without BMI adjustment, R190Q and E288G led to increases in HbA1c (Supplementary Figure 7b). The rarer R101H variant (MAF=0.02%, up to 227 carriers) showed a nominally significant association with lower BMI (P=0.036) but not with HbA1c (P=0.15).

Overall, these data highlight that LoF *GIPR* variants decrease body weight but increase glycaemia, and the latter effect is more pronounced after accounting for changes in BMI. Furthermore, despite conferring increased cAMP response to GIP, the common E354Q variant shows phenotypic consequences similar to the other LoF variants.

### 2.8 Similar impacts of GIP and tirzepatide on *GIPR* variants signalling responses

GIPR agonism is an important component of tirzepatide action (47, 48), so we next investigated how *GIPR* coding variation might influence responses to this agent. Focussing on the ten highest prevalence variants from gnomAD, we tested cAMP concentration responses to GIP or tirzepatide stimulation in AD293 cells (Figure 8a-d). Using the ΔLogE_max_/EC_50_ method to quantify variant effects (49), E354Q showed increased cAMP signalling, whereas E288G, R190Q and R101H showed markedly reduced responses to both ligands (Figure 8b). Statistically significant differences between GIP and tirzepatide were observed with R101H and R190Q variants, the importance of which is unclear as both ligands were very weak agonists in both cases. S64A, R136W and R217L responses were moderately reduced compared to wild-type GIPR for both GIP and tirzepatide (Figure 8c, 8d). Selected variant cAMP signalling effects were independently confirmed using untagged GIPRs (Supplementary Table 5), with relative potency shifts similar to those seen with SNAP-tagged GIPR in Figure 8.

**Figure 8.**
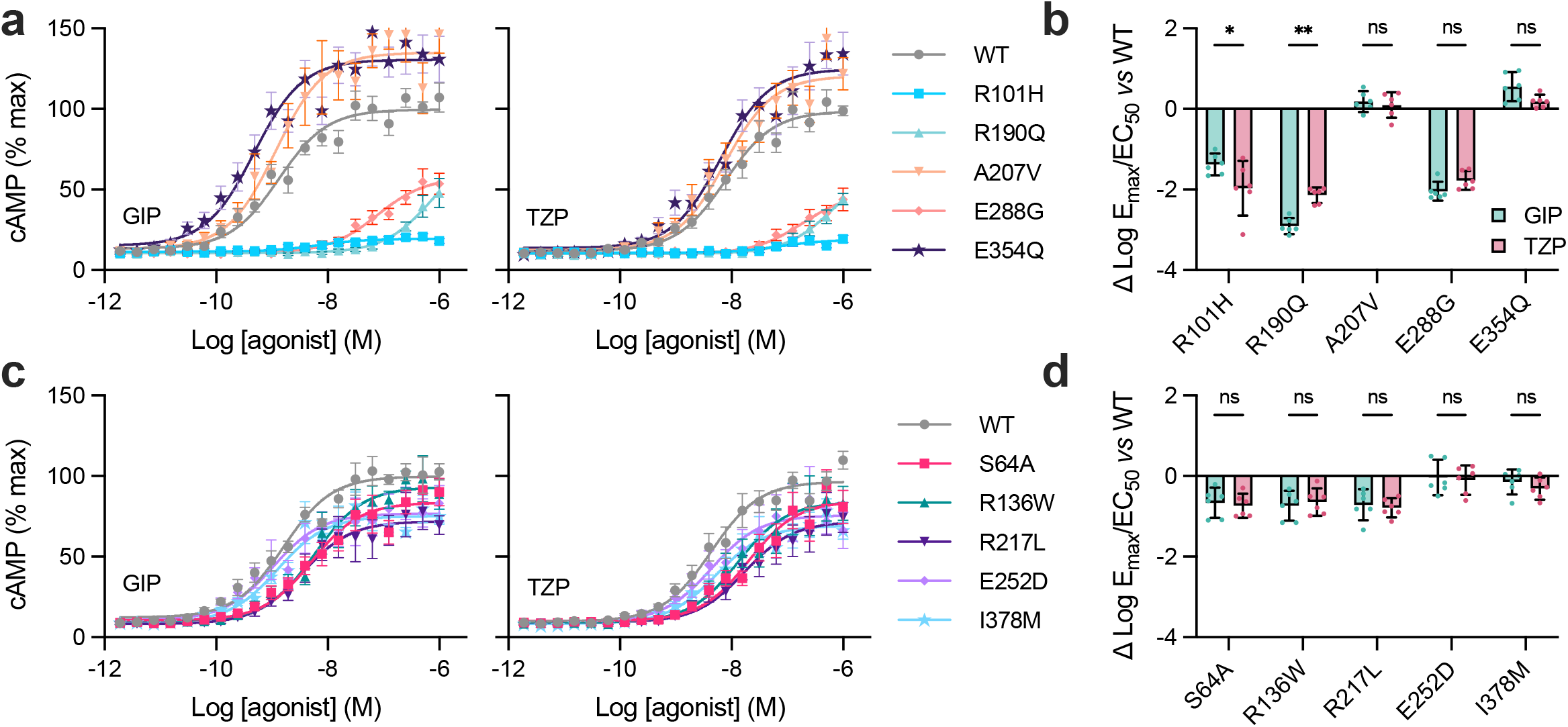
cAMP signalling with tirzepatide. (**a**) cAMP responses in AD293 cells expressing wild-type or variant SNAP-GIPR in response to GIP(1-42) or tirzepatide (TZP), *n*=6. Assays were performed in the presence of 0.1% BSA. (**b**) Variant effect on cAMP signalling analysed using the ΔLogE_max_/EC_50_ method, with statistical comparison of GIP *versus* tirzepatide effect by two-way matched ANOVA with Sidak’s test. (**c**) Same as (a) but different selection of variants. (**d**) Same as (b) but different variants. *p<0.05, **p<0.0001, using indicated statistical test. All data represented as mean ± SEM, except for ΔLogE_max_/EC_50_ data, which shows 95% confidence intervals to highlight differences for variant compared to wild-type; individual data points are shown in some cases.

Thus, across the tested *GIPR* variants, the functional disruptions in cAMP responses to tirzepatide appear to be similar to those in response to GIP(1-42).

## 3 Discussion

In this paper, we have performed a detailed characterisation of 30 *GIPR* missense variants, including pharmacological, cell biological, structural and genomic effects in both AD293 and INS-1 β-cells. We found that a substantial proportion of missense mutations led to significant changes in cAMP signalling, driven jointly by impaired surface delivery of mature receptor to the plasma membrane and intrinsic functional defects. Our structural analysis supports a model in which multiple GIPR variants destabilize the active conformation through disruption of a conserved TM6– TM4 network, providing mechanistic insight into how distal mutations compromise GIP signalling. Leveraging human biobank genetic data allowed us to establish phenotypic correlates of altered GIPR function, which confirmed the reported effect of *GIPR* LoF to lower body weight but also robustly identified effects of *GIPR* LoF on increased glycaemia and T2D risk.

Interest in understanding the effect of coding mutations on GPCR function has increased in recent years, aided by the wide availability of exome sequencing data. Important information has been gleaned from coding variants predicted to be highly deleterious, e.g. due to frameshifts, splice site mutations or premature truncations, as such mutations are very likely to produce a null receptor phenotype (15). However, such mutations are usually very rare. In contrast, more subtle missense variants account for a larger proportion of genomic variation, but their effects are often not straightforward to predict, even with the generation of AI-based tools such as AlphaMissense (50). Experimental studies are often required to classify variants to delineate their phenotypic effects by combining functionally similar variants into groups to increase statistical power. Our functional characterisations led to far clearer phenotypic associations than those achieved by in silico bioinformatic predictions, for example, compared to reported minimum P-values for predicted *GIPR* masks for BMI (P=7.0×10^−4^) and HbA1c (P=3.5×10^−3^) in the same UKBB population (500k WGS v2) (51).

Missense variants in the class B1 GPCR family have attracted recent attention, with a series of recent studies focussed on *GLP1R, GCGR* and *GIPR* (19–21, 52, 53). Each of these studies have highlighted altered surface expression of missense variants as a key driver to altered function. In the present work we confirmed that many *GIPR* missense variants fail to express at adequate levels at the plasma membrane, and we provide insights into the reasons for these changes. In line with a recent study (30) evaluating a wide range of genes, our results point to reduced protein stability leading to degradation during biosynthesis, rather than defects associated with mistargeting to different intracellular locations or accelerated constitutive internalisation/turnover leading to increased lysosomal degradation as a major underlying driver for the reduced levels of missense GIPR variants at the plasma membrane. A prominent role for β-arrestin-driven endosomal signalling in mediating the effects of *GIPR* variants was recently reported (21), which has not been examined in our study. We did however evaluate GIPR internalisation, for which the agonist-dependent fraction is thought to be at least in part β-arrestin dependent (54, 55). The wild-type GIPR shows a high constitutive internalisation rate which is only moderately increased in response to agonist and, with EC_50_ for steady state internalisation (which measures receptors retained intracellularly after one hour of vehicle or agonist exposure but does not account for individual receptors cycling in and out of the endocytic compartment) measured in the tens of nM, it is challenging to study these processes at physiologically relevant GIP concentrations in the pM range. It is plausible that, in addition to the small degree of GIP-induced potentiation of intracellular GIPR localisation, a proportion of GIP/GIPR complexes might also enter the endocytic pathway via constitutive receptor internalisation, without requiring β-arrestins, but still being available to contribute to signalling from intracellular compartments. Because our internalisation assays directly read out the amount of internalised receptor, as well as the percentage, we could easily observe how large between-variant differences in surface expression are a major limiting factor to the achievable total amount of GIPR that can be delivered to endosomes. Further work is needed to understand the precise nature and downstream impacts of both wild-type and variant GIPR signalling at diverse intracellular compartments, as well as effects of acute over prolonged periods of vehicle, GIP or long-acting GIPR agonist stimulation.

Our integrative simulation analysis provides a structural framework for understanding how naturally occurring GIPR variants contribute to altered receptor function. Although the prioritised mutations do not directly contact the GIP peptide, each induces distinct perturbations in inter- and intra-helical interactions within the transmembrane region, with consequences for receptor stability and signalling. Notably, the R190Q variant disrupts local TM2 interactions and long-range stabilising contacts with TM1 and TM6, correlating with reduced surface expression and signalling output. Similarly, E288G, while structurally reminiscent of the wild-type apo state, perturbs critical intra-TM2 interactions and alters peptide positioning, consistent with its partial functional rescue. The E354Q variant impairs a key TM6–TM4 inter-helical contact, leading to a subtle yet functionally relevant shift in the TM6 kink angle distribution toward a less active conformation. This effect on receptor dynamics highlights the sensitivity of the TM6–TM4 interface in maintaining active-state geometry. Finally, the modest but reproducible long-range perturbations observed in the extracellular R101H variant suggest that surface-proximal residues can allosterically modulate transmembrane packing.

In the context of significant interest in GIPR antagonism as a therapeutic approach for obesity, our results from LoF *GIPR* variants may shed light on potential metabolic effects of this strategy. Whilst the weight-lowering effects of LoF *GIPR* variants are now established (14–16, 21), their glycaemic effects have been less clear. Whilst the common E354Q variant was already known to confer mild impairments in glucose tolerance (13), the unusual receptor pharmacology of this variant has limited its confident categorisation as LoF. Unlike other LoF variants, we found that E354Q confers increased cAMP response to GIP and increased constitutive insulin secretion. However, by grouping functionally similar variants, we find that E354Q confers robust phenotypic associations with BMI, HbA1c and T2D, similar to the other *GIPR* LoF variants. Our observed effects of *GIPR* LoF on increased HbA1c and T2D risk contrast with the expected glucose-lowering effects of reduced body weight. One explanation for these paradoxical observations is that reduced β-cell GIPR activity leads to impaired insulin secretion. Alternatively, with increasing evidence that GIPR plays a role in glucose uptake by adipocytes (47, 56, 57), GIPR LoF might impair insulin sensitivity or some other aspect of adipocyte biology to modulate whole body glucose homeostasis. Whilst genetic approaches model the impact of lifelong changes to GIPR function, the possibility that their metabolic impacts are established early in life cannot be excluded. The relative merits of GIPR agonism *versus* antagonism in humans remain unresolved, with a need for studies using pharmacologically optimised GIP-specific ligands rather than GLP-1/GIP combinations to provide a clearer picture of the overall effects on body weight control and glucose homeostasis (58).

## 4 Materials and methods

### 4.1 Peptides

GIP(1-42) and tirzepatide were obtained from WuXi AppTec, China.

### 4.2 Plasmids and adenoviruses

Wild-type and single missense variant SNAP-GIPR plasmid constructs, in which the endogenous GIPR signal peptide was removed and replaced with a SNAP_f_ tag with upstream signal peptide, were obtained from Genewiz, UK. Halo-GIPR was generated in house as previously described (43). Adenoviral SNAP-tagged human GIPR wild-type or variant constructs were generated by VectorBuilder.

### 4.3 Cell culture

Adherent HEK293 (AD293) were cultured in DMEM supplemented with 10% foetal bovine serum (FBS) and 1% penicillin/streptomycin. INS-1 832/3 cells with CRISPR/Cas9 deletion of the endogenous *Gipr* gene were a gift from Dr Jacqueline Naylor (AstraZeneca) (28), referred to herein as “INS-1 cells”, and were cultured in RPMI with 10% FBS and 1% penicillin/streptomycin.

### 4.4 Islet isolation and culture

For *ex vivo* islet experiments, pancreatic islets were isolated from appropriate wild-type or *Gipr* ^-/-^ mice as previously described (59). Pancreata were infused via the common bile duct with RPMI 1640 medium (R8758, Sigma-Aldrich) containing collagenase (1 mg/ml) from *Clostridium histolyticum* (S1745602, Nordmark Biochemicals), dissected, and incubated in a water bath at 37^°^ C for 10 minutes. Islets were subsequently washed and purified using a Histopaque gradient (Histopaque-1119, 11191, Sigma-Aldrich; and Histopaque-1083, 10831, Sigma-Aldrich). Isolated islets were allowed to recover overnight at 37^°^ C in 5% CO_2_ in RPMI 1640 supplemented with 10% (v/v) fetal bovine serum (FBS) (F7524, Sigma-Aldrich) and 1% (v/v) penicillin/streptomycin (P/S) solution (15070-063, Invitrogen).

### 4.5 Assessment of surface GIPR expression by SNAP-tag labelling in cell lines

24 hours before the experiment, AD293 or INS-1 cells were reverse-transfected with *GIPR* variant or wild-type plasmids using Lipofectamine 2000 in 96-well, clear bottomed, black microplates pre-coated with poly-D-lysine. Cells were then labelled using the cell impermeant probe SNAP-Surface AlexaFluor 647 (500 nM, New England Biolabs, USA) for 10 minutes at room temperature to label only surface GIPR. After washing three times with PBS, cells underwent phase contrast and epifluorescence imaging in Krebs-Ringer bicarbonate-HEPES (KRBH) buffer (140 mM NaCl, 3.6 mM KCl, 1.5 mM CaCl_2_, 0.5 mM MgSO_4_, 0.5 mM NaH_2_PO_4_, 2 mM NaHCO_3_, 10 mM HEPES, saturated with 95% O_2_/5% CO_2_; pH 7.4) using an automated widefield microscope (Nikon Ti2E) with computer-controlled stage and focus lock. Several fields-of-view (FoVs) were acquired within each well using a 20X, 0.75 NA air objective. Surface GIPR fluorescence was quantified after flat-field correction using BaSiC (60) and segmenting of cell-containing regions using Phantast (61). Non-specific fluorescence from separate non-transfected cells was subtracted. Data were normalised by dividing by the average specific signal across each experiment, or by the wild-type value, as indicated in the figure legends.

### 4.6 Radioligand binding assay

Radioligand binding to the GIPR variant receptors was performed using the assay procedure described in (62) with some slight modification in the buffer. GIPR membrane protein and wheatgerm agglutinin polyvinyl toluene scintillation proximity assay beads (WGA-PVT SPA beads, Revvity Cat# RPNQ0001) were incubated with a concentration-response curve of [^127^I]Tyr^10^-GIP(1-42)OH (CPC Scientific, San Jose, CA) and [^125^I]GIP(1-42)OH (Revvity, Waltham MA) in 1.0 mM CaCl_2_ (Sigma Cat# 21115), 2.5 mM MgCl_2_ (Boston BioProducts Cat# BM670), 0.003% (v/v) Tween-20 (Roche Cat 11332465001), 0.1% (w/v) Bacitracin (Thermo Scientific Chemicals Cat# J62432-14), 0.1% fatty acid free human serum albumin (Sigma Cat# A3782), 25 mM HEPES (Gibco Cat# 15630-080) pH to 7.4 with KOH, in a 200 µL assay volume. Human GIPR wild type, E354Q, E252D, and A207V used 6 µg of membrane protein, 0.25 mg WGA-PVT SPA bead and ∼0.1 nM [^125^I]GIP(1-42)OH. Human GIPR E288G, E288G+E354Q, and R101H used 12 µg of membrane protein, 0.50 mg WGA-PVT SPA bead and ∼0.3 nM [^125^I]GIP(1-42)OH. The assay was incubated overnight at 25^°^ C, then centrifuged at 1,000 rpm (Beckman Avanti J-15R). Radioligand bound to the GIPR expressing membrane (i.e., in close proximity to the WGA-PVT SPA bead) was detected using a PerkinElmer 2450 MicroPlate Counter. There was no significant specific binding detected in membranes prepared from un-transfected HEK293 cells tested using either condition. Affinity and expression density values were obtained from homologous competition analysis of human [^127^I]Tyr^10^-GIP(1-42)OH versus human [^125^I]GIP(1-42)OH using the competitive binding equation one site homologous (GraphPad Prism version 10.4.1 for Windows).

### 4.7 Assessment of total GIPR expression by SNAP-tag labelling

Experiments were performed and quantified as in Section 4.5, except cells were labelled in parallel (separate wells) using 500 nM SNAP-Surface AlexaFluor 647 or BG-OG (29) for 10 minutes at room temperature.

### 4.8 Immunoblotting for total SNAP-GIPR

INS-1 cells were seeded in a 6-well plate and transfected with 2 µg *GIPR* variant or wild-type plasmids using Lipofectamine 2000 (Thermo Fisher) per the manufacturer’s instructions. 24 hours post transfection the cells were lysed with 1X TNE lysis buffer (20 mM Tris, 150 mM NaCl, 1 mM EDTA, 1% NP40, protease and phosphatase inhibitor cocktails) for 10 minutes at 4^°^ C followed by cell scraping and sonication (3X, 10 seconds each). The lysates were then centrifuged at 12,000 g for 10 minutes at 4^°^ C. The supernatants were collected, fractionated by SDS-PAGE in urea loading buffer (200 mM Tris HCl pH 6.8, 5% w/v SDS, 8 M urea, 100 mM DTT, 0.02% w/v bromophenol blue) and analysed by Western blotting. SNAP-GIPR was detected with an anti-SNAP-tag rabbit polyclonal antibody (P9310S, New England Biolabs, RRID: AB_10631145, 1/1,000) followed by goat anti-rabbit HRP secondary (ab6721, Abcam, RRID: AB_955447, 1/2,000). Post-stripping, tubulin was labelled with anti-α-tubulin mouse monoclonal antibody (T5168, Sigma, RRID: AB_477579, 1/5,000) followed by sheep anti-mouse HRP secondary antibody (ab6808, Abcam, RRID: AB_955441, 1/5,000). Blots were developed with the Clarity Western enhanced chemiluminescence (ECL) substrate system (BioRad) with the ChemiDoc Imaging System (BioRad) and specific band densities quantified in Fiji.

### 4.9 Prediction of variant thermal stability

Experimental GIPR structures (7RA3, 7RBT, 7DTY) were obtained from PDB, and a Alphafold 3-predicted full-length GIPR structure (AF-P48546-F1-model_v4) was obtained from the AlphaFold server (https://alphafold.com). After removing non-GIPR chains from the experimental structures, mCSM-membrane (34) was used to predict the thermostability effect of GIPR mutations present in the structures (the receptor C-terminus is not included in experimental structures) as ΔΔG.

### 4.10 Proteasomal inhibition

INS-1 cells were reverse-transfected with *GIPR* variant or wild-type plasmids in 96-well, clear bottomed, black microplates pre-coated with poly-D-lysine. 12 hours post transfection, the cells were treated with either vehicle or Proteasome Inhibitor I (Merck) for 4 hours. Following this, the cells were labelled with the cell-permeable SNAP-tag probe BG-OG (29) for 10 minutes at 37°C to label total SNAP. Cells were then imaged as in Section 4.5 to establish total GIPR labelling (surface and intracellular). Images were analysed as in Section 4.5.

### 4.11 Constitutive and agonist-mediated internalisation measured by reversible SNAP-labelling

24 hours before the experiment, AD293 or INS-1 cells were reverse-transfected with *GIPR* variant or wild-type plasmids using Lipofectamine 2000 in 96-well, clear bottomed, black microplates pre-coated with poly-D-lysine. Cells were then labelled using the cleavable, cell impermeant probe BG-SS-649 (a gift from New England Biolabs, USA) for 10 minutes at room temperature. Cells were then incubated for 60 minutes in serum-free medium containing 0.1% BSA at 37^°^ C, with or without 100 nM GIP. Media was then removed, cells washed with PBS and then placed in TNE buffer pH 8.6 for imaging. Cells were imaged as in Section 4.5 to establish total surface GIPR labelling, and after each well had been imaged, the cell-impermeant reducing agent Mesna (200 mM) was added to remove any surface BG-SS-649 bound to non-internalised GIPR. After 10 minutes, the well was reimaged again to detect remaining fluorescence from internalised GIPR. Images were analysed as in Section 4.5, with the residual specific fluorescence after Mesna treatment expressed relative to the pre-Mesna value to establish percentage internalisation.

### 4.12 Bafilomycin A1 experiments

24 hours before the experiment, AD293 or INS-1 cells were reverse-transfected with *GIPR* variant or wild-type plasmids using Lipofectamine 2000 in 96-well, clear bottomed, black microplates pre-coated with poly-D-lysine. For AD293 cells, after 12 hours, 100 nM Bafilomycin A1 or vehicle were added to the wells. A further 12 hours later, cells were labelled using SNAP-Surface AlexaFluor 647 and imaged as described in Section 4.5. For INS-1 cells, after 12 hours, 400 nM Bafilomycin A1 or vehicle were added to the wells. After 4 hours of incubation with vehicle of Bafilomycin A1, cells were labelled with SNAP-Surface AlexaFluor 647 and imaged as described in Section 4.5.

### 4.13 Assessing for dominant negative effects by SNAP- and Halo-tag GIPR co-expression

24 hours before the experiment, AD293 cells were reverse-co-transfected with SNAP-tagged *GIPR* variant or wild-type plasmids plus Halo-tagged wild-type GIPR in a 6:1 ratio using Lipofectamine 2000 in 96-well, clear bottomed, black microplates pre-coated with poly-D-lysine. After 24 hours, cells were dual-labelled with SNAP-Surface 488 (0.5 µM, New England Biolabs) and Halo-AlexaFluor 660 (0.5 µM, Promega, UK) for 10 minutes at 37^°^ C. After washing, cells were imaged as in Section 4.5, and surface expression of SNAP- and Halo-tagged GIPR were quantified as in Section 4.5.

### 4.14 Cyclic AMP biochemical measurements

#### Single concentration experiment

AD293 cells were reverse-transfected with *GIPR* variant or wild-type plasmids using Lipofectamine 2000 in 96-well microplates. After 24 hours, cells were treated with GIP (100 pM or 100 nM) or 500 µM isobutylmethylxanthine (IBMX) in serum-free medium containing 0.1% BSA for 30 minutes at 37^°^ C, before terminating the reaction by adding lysis buffer and measuring cellular cAMP by HTRF (cAMP Dynamic 2 assay, Revvity). IBMX was not used in the assays using GIP. cAMP concentration measured from untransfected cells was subtracted to isolate the receptor/agonist-specific response.

#### Concentration response experiments (Imperial College London)

24 hours before the experiment, AD293 cells were reverse-transfected with *GIPR* variant or wild-type plasmids using Lipofectamine 2000 in 12-well plates. Cells were then treated in suspension with a range of concentrations of GIP or tirzepatide in serum-free medium containing 0.1% BSA and 250 µM IBMX for 30 minutes at 37^°^ C, before terminating the reaction by adding lysis buffer and measuring cellular cAMP by HTRF. Data were fit using a 3-parameter logistic model using GraphPad Prism 10.

#### Concentration response experiments (Eli Lilly)

HEK293 cells transiently expressing the human GIPR (NP_000155) or the variants described herein were evaluated by measuring the accumulation of intracellular cAMP using the Gs Dynamic Assay (PerkinElmer), as previously described(62). The assay medium was DMEM (Thermo Fisher Scientific] containing 2 mM Glutamax (Thermo Fisher Scientific), 20 mM HEPES, 0.1% w/v bovine casein (MilliporeSigma), and 250 μM IBMX. Cells were added to treatment plates and incubated at 37^°^ C for 30 minutes and then lysed in the presence of d2-labeled cAMP competitor conjugate and cryptate-conjugated detection antibody. Time-resolved fluorescence was measured with a Pherastar FSX reader (BMG Labtech) and data were analyzed by standard ratio methods. Normalized values were fit to the 4-parameter logistic model using GraphPad Prism 7 software.

### 4.15 Cyclic AMP imaging

AD293 cells were reverse-transfected with *GIPR* variant or wild-type plasmids using Lipofectamine 2000, and co-transduced with the cADDis Green Up BacMam (10 µL/well, Montana Molecular, USA), in 96-well, poly-D-lysine coated microplates. After 24 hours, cells were labelled using 500 nM SNAP-Surface AlexaFluor 647 for 10 minutes at room temperature. Cells were washed three times and then imaged in KRBH buffer containing 6 mM glucose and 0.1% BSA. Imaging was performed at 37^°^ C by wide-field microscopy using the same microscope as in Section 4.5, with 9 fields-of-view for both cADDis and SNAP-Surface fluorescence acquired from each well using a 10X, 0.45 NA air objective. For steady state assays, images were acquired at baseline, 10 minutes after addition of 100 pM or 100 nM GIP, and again 10 minutes after addition of 500 µM IBMX + 50 µM forskolin (FSK). Flatfield correction was performed using BaSiC (60), drift correction using SIFT, and a maximum intensity projection of the cADDis channel was used to segment individual cells. cAMP responses were determined by quantifying cADDis fluorescence intensity for each cell at each time-point. Non-viable cells or non-cell objects were excluded according to criteria for allowable cell IBMX/FSK responses. The GIP effect for each cell was quantified by double normalising cADDis signal to baseline and then to IBMX/FSK. The cADDis mask was then used to quantify SNAP-Surface signal from each cell. Wild-type SNAP-Surface readings were split into deciles, and these deciles were subsequently used to categorise variant-transfected cells to match against wild-type. After trimming the top and bottom deciles, the mean was taken of each decile mean for surface expression level or cAMP response, which achieved matched expression levels by effectively applying an inverse weighting to counteract differences in cell number in each decile for different variants. Ensemble expression levels or cAMP responses were calculated without regard to deciles.

### 4.16 Molecular dynamics simulations

For full methodological details, see Supplementary Methods.

### 4.17 Assessment of surface GIPR expression by SNAP-tag labelling in pancreatic islets

Islets were purified from *Gipr*^-/-^ mice as described in Section 4.4. Islets were transduced with adenoviral wild-type or variant SNAP-tagged *GIPR* constructs (VectorBuilder) per the manufacturer’s instructions. 24 hours post transduction, islets were labelled with SNAP-Surface AlexaFluor 647 for 15 minutes at 37^°^ C. Islets were imaged on a Zeiss LSM-780 inverted confocal microscope fitted with a 63X/1.4 NA oil immersion objective. Receptor surface levels were quantified in Fiji using a macro employing full width at half maximum (FWHM) designed by the Facility for Imaging and Light Microscopy (FILM) at Imperial College London and expressed relative to wild-type GIPR.

### 4.18 Hormone secretion assays from pancreatic islets

Isolated islets used for insulin secretion assays were treated in 1.5 mL microcentrifuge tubes. Eight-ten islets were used per well with two-three technical replicates per condition. Islets were preincubated for 1 hour in Krebs-Ringer bicarbonate–Hepes (KRBH) buffer (140 mM NaCl, 3.6 mM KCl, 1.5 mM CaCl_2_, 0.5 mM MgSO_4_, 0.5 mM NaH_2_PO_4_, 2 mM NaHCO_3_, 10 mM Hepes, saturated with 95% O_2_/5% CO_2_; pH 7.4) containing 0.1% (w/v) bovine serum albumin (BSA) and 3 mM or 11 mM glucose for insulin or glucagon secretion, respectively. This was followed by incubation with 11 mM or 0.5 mM glucose ± 100 nM GIP in KRBH buffer for insulin or glucagon secretion, respectively, in a shaking 37^°^ C water bath (80 rpm) for 30 minutes. At the end of the treatment, supernatants containing the secreted insulin or glucagon were collected, centrifuged at 1,000 g for 5 minutes, and transferred to fresh tubes. To determine total insulin or glucagon contents, islets were lysed using acidic ethanol [75% (v/v) ethanol and 1.5 mM HCl]. The lysates were sonicated 3 × 10 seconds in a water bath and centrifuged at 10,000 g for 10 minutes, and the supernatants collected. The samples were stored at -20^°^ C until the insulin or glucagon concentrations were determined using an Insulin Ultra-Sensitive HTRF Assay kit (62IN2PEG, Revvity) or an HTRF Glucagon Detection Kit (62CGLPEG, Revvity) respectively, according to the manufacturer’s instructions. GraphPad Prism 9.0 was used for the generation of the standard curve and sample concentration extrapolation. The total insulin or glucagon content was calculated by adding the secreted insulin to the insulin content of the lysates.

### 4.19 Statistics for wet laboratory experiments

The average of technical replicates was considered as one biological replicate for in vitro experiments. Experiments were performed with all conditions tested in parallel to allow matched analyses, which were performed using Prism 10.5.1 (GraphPad Software). Specific statistical tests, including corrections for multiple comparisons, are indicated in the figure legends. Statistical significance was inferred when p<0.05. Data are represented as mean ± SEM, or with 95% confidence intervals where specifically indicated.

### 4.20 Exome-wide association study: ethics and study approval

The UK Biobank operates under approval from the North West Multi-centre Research Ethics Committee (REC reference 13/NW/0157) as a Research Tissue Bank (RTB), with informed consent obtained from all participants. This approval allows researchers to conduct studies without requiring separate ethical clearance, as they are covered by the RTB approval. Initially granted in 2011, this approval is renewed every five years, with successful renewals completed in 2016 and 2021. The work described for this analysis was approved by the UK Biobank through resource application number 9905.

### 4.21 Exome-wide association study: data, quality control and analysis

For full methodological details, see Supplementary Methods.

### 4.22 Data availability

All experimental data from wet laboratory experiments are included in the Supporting Data Values file. The UKBB phenotype and WES data used in this study are accessible to registered researchers via the UKBB data access protocol. Details on how to register for data access can be found at https://www.ukbiobank.ac.uk/enable-your-research/apply-for-access. Data for this study were obtained under Resource Application Number: 9905.

## 5 Acknowledgements

This work was supported by an LRAP award to A.T., B.J. and K.W.S. B.J. is supported by an MRC Clinician Scientist Fellowship (MR/Y00132X/1). The A.T. group is funded by grants from Diabetes UK (19/0006094), the MRC (MR/X021467/1), and the Wellcome Trust (301619/Z/23/Z), the latter two in collaboration with B.J. E.J.G., J.R.B.P., K.K.O. and N.J.W. acknowledge funding from the Medical Research Council (Unit Programmes: MC_UU_00006/1 and MC_UU_00006/2) and the NIHR Cambridge Biomedical Research Centre (NIHR203312). The Section of Endocrinology at Imperial College London is funded by grants from the MRC, NIHR and is supported by the NIHR Biomedical Research Centre Funding Scheme and the NIHR/Imperial Clinical Research Facility. The views expressed are those of the author(s) and not necessarily those of the NHS, the NIHR, or the Department of Health. For the purpose of Open Access, the author has applied a CC BY public copyright licence to any Author Accepted Manuscript version arising from this submission.

## Notes

**Conflict of interest statement** This work was supported by an LRAP award to A.T., B.J. and K.W.S. B.J. has received grant funding and is a consultant for Metsera Inc. A.T. has received funding from Sun Pharmaceuticals. J.R.B.P. and E.J.G. are employees and shareholders of Insmed Inc. R.D., K.K.O. and N.J.W. receive research funding from Eli Lilly. J.R.B.P. receives research funding from GSK.

### Competing Interest Statement

This work was supported by an LRAP award to A.T., B.J. and K.W.S. B.J. has received grant funding and is a consultant for Metsera Inc. A.T. has received funding from Sun Pharmaceuticals. J.R.B.P. and E.J.G. are employees and shareholders of Insmed Inc. R.D., K.K.O. and N.J.W. receive research funding from Eli Lilly. J.R.B.P. receives research funding from GSK.

