## Supplementary Table 1 for "Glycaemic and bodyweight effects of *GIPR* coding variation reflect differences in both surface expression and intrinsic functional impairment"

**Supplementary Table 1**: Human [^125^I]GIP(1-42)OH binding to human GIP-R receptor mutants expressed in HEK-293 cells.

| **Receptor** | **K_d_ ^a^ (nM)**  **(SEM, n)** | **B_max_ ^b^ (fmol/mg protein)**  **± SEM (n)** |
| --- | --- | --- |
| hGIPR WT | 0.383  (0.038, 5) | 1340  ± 280 (5) |
| hGIPR (E354Q) | 0.327  (0.045, 4) | 1150  ± 130 (4) |
| hGIPR (E252D) | 0.422  (0.038, 5) | 1200  ± 210 (5) |
| hGIPR (A207V) | 0.467  (0.069, 3) | 1680  ± 30 (3) |
| hGIPR (E288G) ^c^ | 10.1  (7.5, 5) | 2870  ± 1290 (5) |
| hGIPR (E288G + E354Q) ^c^ | 23.2  (7.6, 5) | 4130  ± 1730 (5) |
| hGIPR (R101H) ^c^ | 2.72  (1.67, 6) | 927  ± 397 (6) |

^a^ K_d_ values are reported as the geometric mean with the SEM and the number of independent experiments in parentheses.

^b^ B_max_ values are the arithmetic mean ± SEM with the number of independent experiments in parentheses.

^c^ Denotes that, though expression was seen, due to the lower affinity for human [^125^I]GIP(1-42)OH at these three mutant receptors an accurate K_d_ and B_max_ were difficult to obtain using this radioligand. The bottom of the fitted [^127^I]Tyr^10^-GIP(1-42)OH curve was used to define nonspecific binding for these three mutant receptors. Nonspecific binding was defined by 100 nM or 1 µM GIP(1-42)OH for the remaining mutant and wild type human GIPR.
