## Supplementary Table 5 for "Glycaemic and bodyweight effects of *GIPR* coding variation reflect differences in both surface expression and intrinsic functional impairment"

**Supplementary Table 5**: hGIP(1-42)NH_2_ and tirzepatide cAMP responses in cells expressing untagged human GIP-R and variants. Assays were performed in the presence of 0.1% casein.

|  | **cAMP**  **Human GIP(1-42)NH_2_**  **in human GIP-R receptor mutants expressed in HEK-293 cells.** | | **cAMP**  **Tirzepatide**  **in human GIP-R receptor mutants expressed in HEK-293 cells.** | |
| --- | --- | --- | --- | --- |
| **Receptor** | **EC_50_ nM**  **(SEM, n)** | **E_max_ %**  **(SEM, n)** | **EC_50_ nM**  **(SEM, n)** | **E_max_ %**  **(SEM, n)** |
| hGIPR WT | 0.00151 (.0001,4) | 96.4 (0.82, 4) | 0.00148 (0.0002, 4) | 90.6  (0.89, 4) |
| hGIPR (E354Q) | 0.000383 (0.00003, 2) | 98.7 (0.275, 2) | 0.000821 (0.0001, 2) | 104  (2.50, 2) |
| hGIPR (E252D) | 0.00189 (0.0002, 2) | 104 (1.15, 2) | 0.00197 (0.0001, 2) | 97.1  (5.18, 2) |
| hGIPR (A207V) | 0.00160 (0.0002, 2) | 106 (9.98, 2) | 0.00180 (0.0003, 2) | 94.5  (2.86, 2) |
| hGIPR (E288G) ^c^ | 0.338 (0.0592, 2) | 101 (2.63, 2) | 0.191 (0.0194, 2) | 101  (8.69, 2) |
| hGIPR (E288G + E354Q) ^c^ | 0.726 (0.0719, 2) | 97.1 (2.09, 2) | 0.572 (0.0575, 2) | 98.1  (2.92, 2) |
| hGIPR (R101H) ^c^ | 0.125 (n/a, 1) | 100  (n/a, 1) | 0.259 (n/a, 1) | 97.1  (n/a, 1) |
