## Supplementary Methods for "Glycaemic and bodyweight effects of *GIPR* coding variation reflect differences in both surface expression and intrinsic functional impairment"

### 1 Molecular dynamics simulations

System Preparation: The membrane-protein system was prepared using a custom Desmond System Builder file optimized for OPM-aligned membrane-bound proteins. The protein-lipid system was embedded in a pre-equilibrated POPC bilayer with 15 Å buffers in each dimension to ensure adequate solvation and spatial separation. The SPC water model was used to solvate the system, and a 0.15 M NaCl concentration was added to approximate physiological ionic strength. Counter-ions were included to neutralize the system. Overlapping crystallographic water molecules were removed, and solvent van der Waals radii were scaled by 0.75 to reduce steric clashes. The OPLS4-based S-OPLS force field was assigned to all system components. Hydrogen mass repartitioning was enabled to support the use of larger integration time steps in subsequent MD simulations. Non-crystallographic water molecules were assigned to an energy group to enable selective control during membrane relaxation and equilibration. System construction was performed using Schrodinger’s Desmond preparation framework.

All molecular dynamics (MD) simulations were performed under periodic boundary conditions with temperature maintained at 300 K unless otherwise stated using the Desmond simulation engine (1) within the (2) software suite. A custom membrane protein equilibration and relaxation protocol was employed, adapted from a procedure originally developed by Lupyan and colleagues (unpublished, Schrödinger Inc., 2009). This protocol is designed to gradually relax steric clashes and artifacts associated with protein crystallography and membrane packing, especially following system assembly in a lipid bilayer environment. The system underwent a staged equilibration process, comprising five distinct phases:

1. Initial Brownian Dynamics (NVT, 10 K, 50 ps): A short Brownian dynamics simulation at 10 K and constant volume (NVT ensemble) was used to minimize high-energy contacts. Solute heavy atoms were harmonically restrained (50 kcal/mol/Å²). A small timestep (0.001–0.003 ps) and the Brownie integrator were used to accommodate potentially large initial forces.
2. High-Pressure Packing (NPT, 100 K, 20 ps): The system was then equilibrated at 100 K and high pressure (1000 atm) using another Brownian dynamics step in the NPT ensemble to enhance membrane-protein packing. Harmonic restraints were applied anisotropically to membrane heavy atoms in the z-dimension and to solute heavy atoms (20 kcal/mol/Å²). A GaussianBarrier plugin was applied to prevent water intrusion between the membrane and protein, implementing two Gaussian repulsive potentials centered at ±10 Å along the z-axis (σ = 0.5 Å, A = 10 kcal/mol) to act only on water molecules previously reassigned to a distinct energy group.
3. Further Packing Under NPgT (100 K, 100 ps): A 100-ps simulation at 100 K and 1000 atm was conducted using the MTK integrator in the NPgT ensemble, allowing anisotropic pressure coupling in x–y versus z. Restraints were weakened (2 kcal/mol/Å² on lipid headgroups and 10 kcal/mol/Å² on solute heavy atoms), and Gaussian barriers were retained.
4. Gradual Heating and Pressure Reduction (NPgT, 100→300 K, 150 ps): The system was then heated from 100 K to 300 K over 150 ps using an annealing schedule. Restraints remained as above, and the barostat and thermostat coupling constants were set to 2.0 and 0.1 ps, respectively. A simplified GaussianForce plugin (σ = 5 Å, A = 2 kcal/mol centered at z = 0) replaced the dual-barrier setup to continue discouraging water intrusion.
5. Production Run (NPgT, 300 K): After equilibration, restraints and Gaussian barriers were removed, and production simulations were run under NPgT conditions at 300 K and 1 atm. Surface tension was maintained in the x–y plane, with anisotropic pressure scaling.A timestep scheme of 2 fs (with multiple timestep RESPA integration for bonded and nonbonded interactions) was used during standard MD runs, except during Brownian dynamics stages. Nose–Hoover thermostats and Martyna–Tobias–Klein barostats were used to maintain temperature and pressure, respectively (3, 4) . Final simulations times were approximately 1 microsecond. Approximately 750 ns of production trajectory data were collected at appropriate intervals for downstream analysis. Protein-protein interactions, kink-angles, and clustering analyses were all performed using in-house custom workflows.

### 2 Exome-wide association study: data and quality control

We accessed whole exome sequence (WES) data on up to 469,835 participants from the UKBB study (470k WES release, July 2022) (5). We analysed data on 465,797 individuals after excluding samples with excess heterozygosity, autosomal variant missingness on genotyping arrays ≥5%, or were not included in the subset of phased samples as defined in Bycroft *et al*. (6). For our European-only sample (n=434,438 individuals), we excluded participants who were not of broadly European genetic ancestry. For each outcome, further exclusions were implemented due to incomplete phenotype data (see Supplementary Table 4). Continuous outcome traits were BMI and HbA1c (excluding diabetes cases). T2D was the only binary outcome (defined in Supplementary Table 4). Outcome data were taken from the first UKBB study visit, unless otherwise specified in Supplementary Table 4. Rare variant burden testing was performed on both the European-only and the full multiethnic UKBB samples. As results were consistent in both strata and statistical power was enhanced in the latter by its larger sample size and by incorporating rare variants present in non-Europeans, single-variant analysis was conducted only on the full multiethnic UKBB sample. All data processing and analyses were performed within the UKBB Research Analysis Platform (RAP; <https://ukbiobank.dnanexus.com/>). This is a cloud-based computing environment and the central repository for UKBB WES and phenotype data. WES data are stored as population-level variant call format (VCF) files, aligned to GRCh38. In addition to the processing applied to the released data, as documented in Backman *et al.* (5), we performed further quality control measures as described in Gardner *et al.* (7) and detailed in the MRC-EPID WES pipeline (<https://github.com/mrcepid-rap/>).

### 3 Single-variant and burden association tests

We conducted burden tests by collapsing rare (MAF < 1%) *GIPR* variants with similar experimental functional consequences, specifically their 100 pM GIP cAMP responses and levels of surface expression in AD293 cells. We assigned variants to reduced, neutral and increased response groups, with “reduced” defined as a statistically significant reduction of at least 50% below that of the wild-type receptor. The common missense variant, E354Q, was excluded from burden tests due to its high population prevalence (> 19% in UKBB) (8). The remaining *GIPR* variants were grouped based on their impact on surface expression into two categories: (a) 11 variants with reduced surface expression, and (b) 15 variants with a neutral impact on expression compared to wild-type. The I378M variant was the only one to exhibit increased surface expression and, therefore, could not be tested in a group for burden analysis due to the low number of carriers. Similarly, *GIPR* variants were also categorized based on their cAMP response to GIP stimulation into three categories: (a) 14 variants with reduced cAMP response, (b) 11 variants with a neutral impact on cAMP response, and (c) 2 variants with increased cAMP response. We performed rare variant burden association testing using two statistical methods: STAAR ("variant-Set Test for Association using Annotation infoRmation") (9) and a generalized linear regression model as implemented in the Python package ‘Statsmodels’. STAAR effectively accounts for population structure and relatedness, so was considered to be particularly applicable to analyses of the full multiethnic sample where population differences are potential confounders. The generalized linear model is widely used but it is less robust for analysing rare variants, especially when population structure and relatedness are significant factors, so was included as a sensitivity measure. We also performed single-variant association testing using BOLT-LMM, a mixed-model algorithm (v2.4.1) (10). BOLT-LMM calculates the genetic relationship matrix (GRM), or kinship matrix, to account for potential confounding by population stratification and hidden relatedness. We applied BOLT-LMM in the customized applet for each outcome, with default settings and the ‘ImmInfOnly’ flag, which limits BOLT-LMM to an “infinitesimal” mixed model. BOLT-LMM requires two main data inputs: (i) genotypes from genotyping arrays, filtered for variants with a minor allele count > 100, to build a null model, and (ii) a broader set of imputed variants for association testing. For the first input, we accessed UKBB array genotype data available on the RAP, in a sample identical to that used for rare variant testing. For the latter, we included all 27,040,378 single markers in the input to BOLT-LMM, regardless of filtering status. Single variant-level BOLT association summary statistics were then extracted for the 30 functionally characterized variants. Significance thresholds were established using Bonferroni correction for multiple testing of 28 variants in the 2 main uncorrelated phenotypes (BMI and HbA1c) (i.e. *P*-value<9x10^-4^ was considered to be statistically significant and it was calculated as 0.05 / [28 variants x 2 phenotypes]) (11). All analyses were performed using bespoke applets designed for the RAP as described in the MRC-EPID WES pipeline (<https://github.com/mrcepid-rap/>), controlled for sex, age, age^2^, the first ten genetic ancestral principal components (PCs) as calculated by Bycroft *et al*. (6) and WES release batch (50k, 200k, 450k).
