## Supplementary Figures for "Glycaemic and bodyweight effects of *GIPR* coding variation reflect differences in both surface expression and intrinsic functional impairment"

**Supplementary Figure 1**

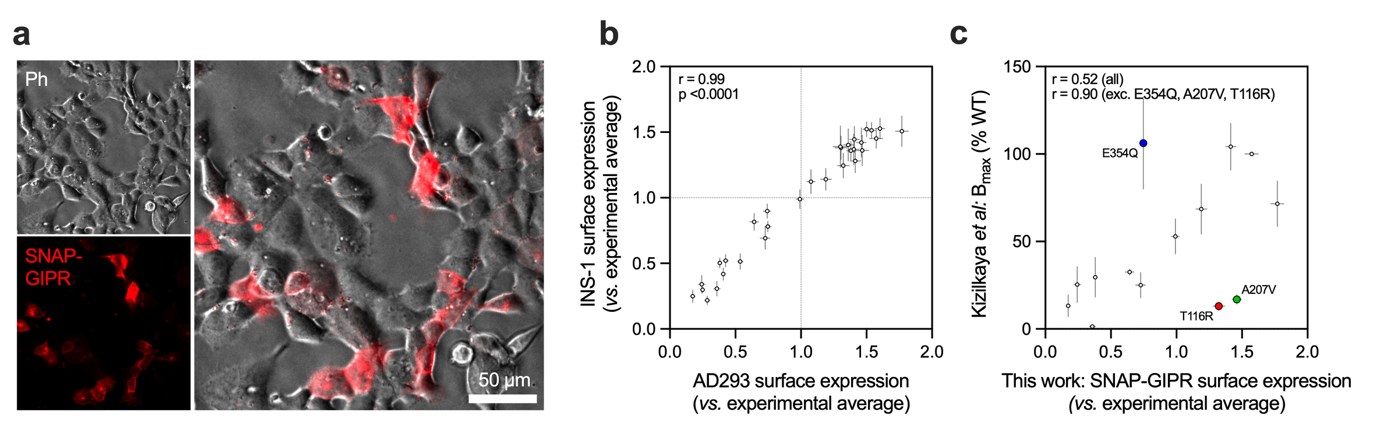

**Supplementary Figure 1.** (**a**) Example image from high content analysis of GIPR surface expression. Scale bar = 50 µm. (**b**) Comparison of GIPR variant surface expression in AD293 and INS-1 cells, using data from Figure 1c and 1d. (**c**) Comparison of AD293 surface expression levels from Figure 1c and the B_max_ for ^125^I-GIP measured by Kizilkaya *et al.* All data represented as mean ± SEM.

**Supplementary Figure 2**

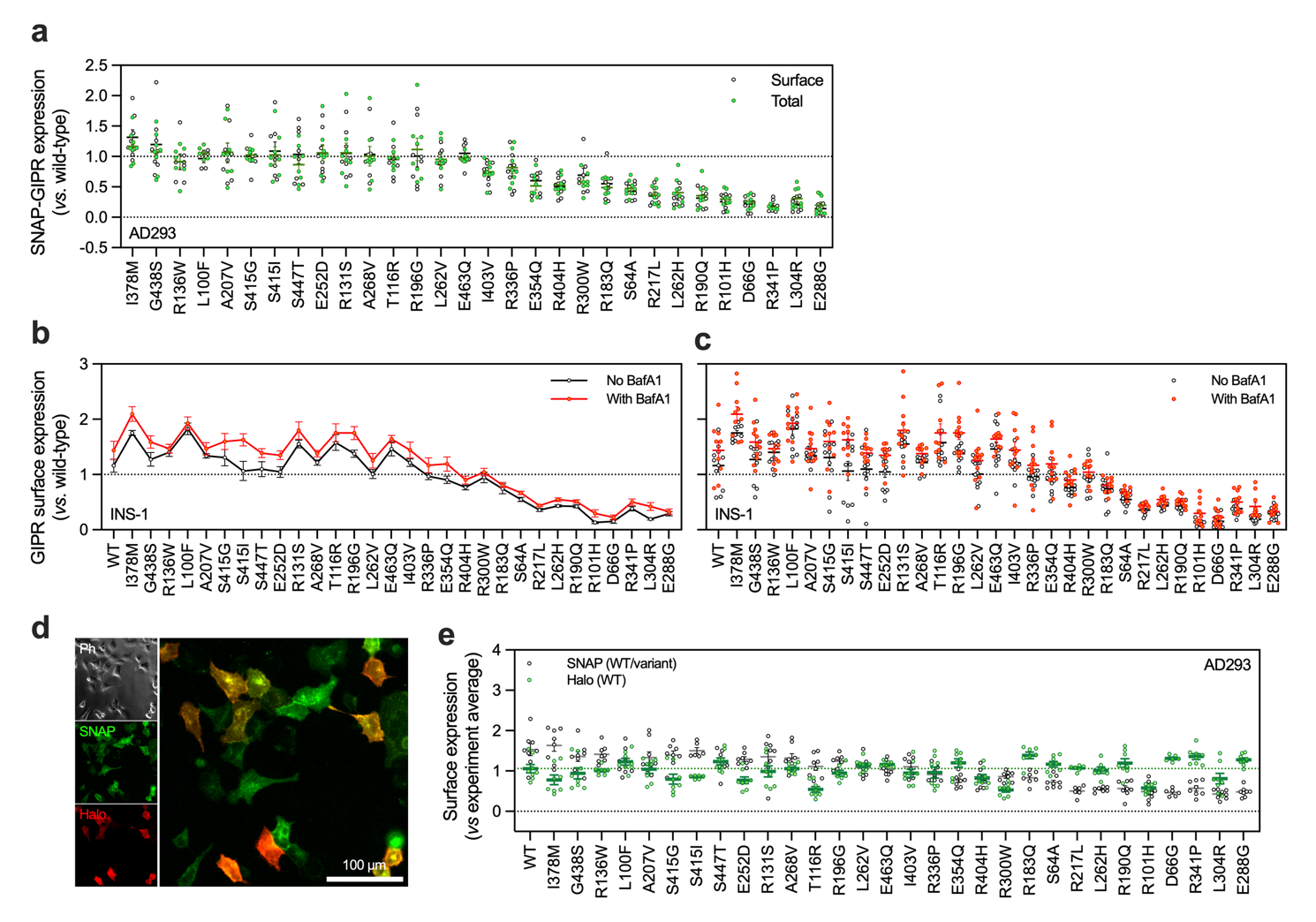

**Supplementary Figure 2.** (**a**) Individual datapoints from Figure 2a, showing surface and total expression of wild-type and variant SNAP-GIPR in AD293 cells, *n*=8, with normalisation to the wild-type (dotted line). (**b**) Effect of bafilomycin A1 treatment (4 hours, 400 mM) on relative surface expression of SNAP-GIPR in AD293 cells, *n*=8, scaled to experimental average without PSI, and compared using two-way matched ANOVA with Sidak’s test with/without bafilomycin A1. (**c**) Individual data points from Supplementary Figure 2b. (**d**) Representative image from high content analysis of SNAP- and Halo-GIPR co-expression with surface-specific dual colour labelling. Scale bar = 100 µm. (**e**) Individual data points from Figure 2f, showing surface expression of wild-type Halo-GIPR and wild-type/variant SNAP-GIPR in AD293 cells, *n*=8, with scaling to the experimental average for each probe.

**Supplementary Figure 3**

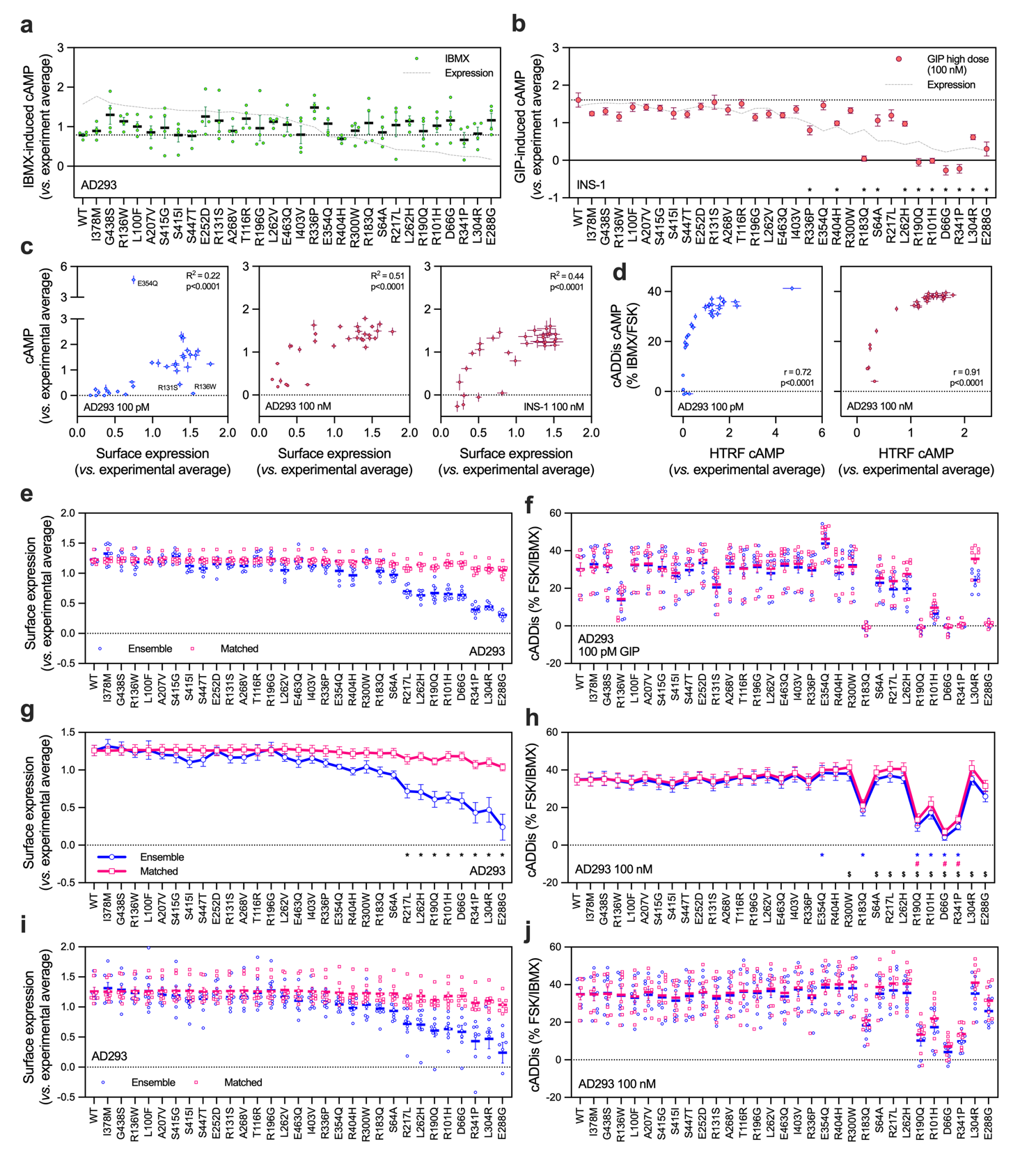

**Supplementary Figure 3.** (**a**) cAMP responses measured by HTRF in the presence of 500 µM IBMX but no GIP in AD293 cells, *n*=4. Results are scaled to the experimental average and compared by two-way matched ANOVA with Dunnett’s test *versus* wild-type. The dotted line shows the wild-type mean and the dashed line shows the surface expression level from Figure 1c. (**b**) As for (a) but showing responses to 100 nM GIP in INS-1 cells, *n*=8. (**c**) The relationship between surface expression levels of wild-type and variant GIPR in AD293 and INS-1 cells (from Figure 1c and 1d) and the corresponding HTRF cAMP response to 100 pM or 100 nM GIP. (**d**) The relationship between HTRF and cADDis biosensor measured cAMP responses. (**e**) Individual data points from Figure 3e, showing surface GIPR expression from ensemble (unselected) and expression-matched analysis, *n*=8, with scaling to the experimental average. (**f**) Individual data points from Figure 3f, showing cADDis response to 100 pM GIP, *n*=8. (**g**) and (**h**) show equivalent data and analysis to data from Figure 3e and 3f, except with higher GIP concentration (100 nM); (**i**) and (**j**) show the individual data points from (g) and (h). *p<0.05 (variant *versus* wild-type from ensemble cells), #p<0.05 (variant *versus* wild-type from expression-matched cells), $p<0.05 (ensemble *versus* expression-matched cells). All data represented as mean ± SEM with individual data points shown in some cases.

**Supplementary Figure 4**

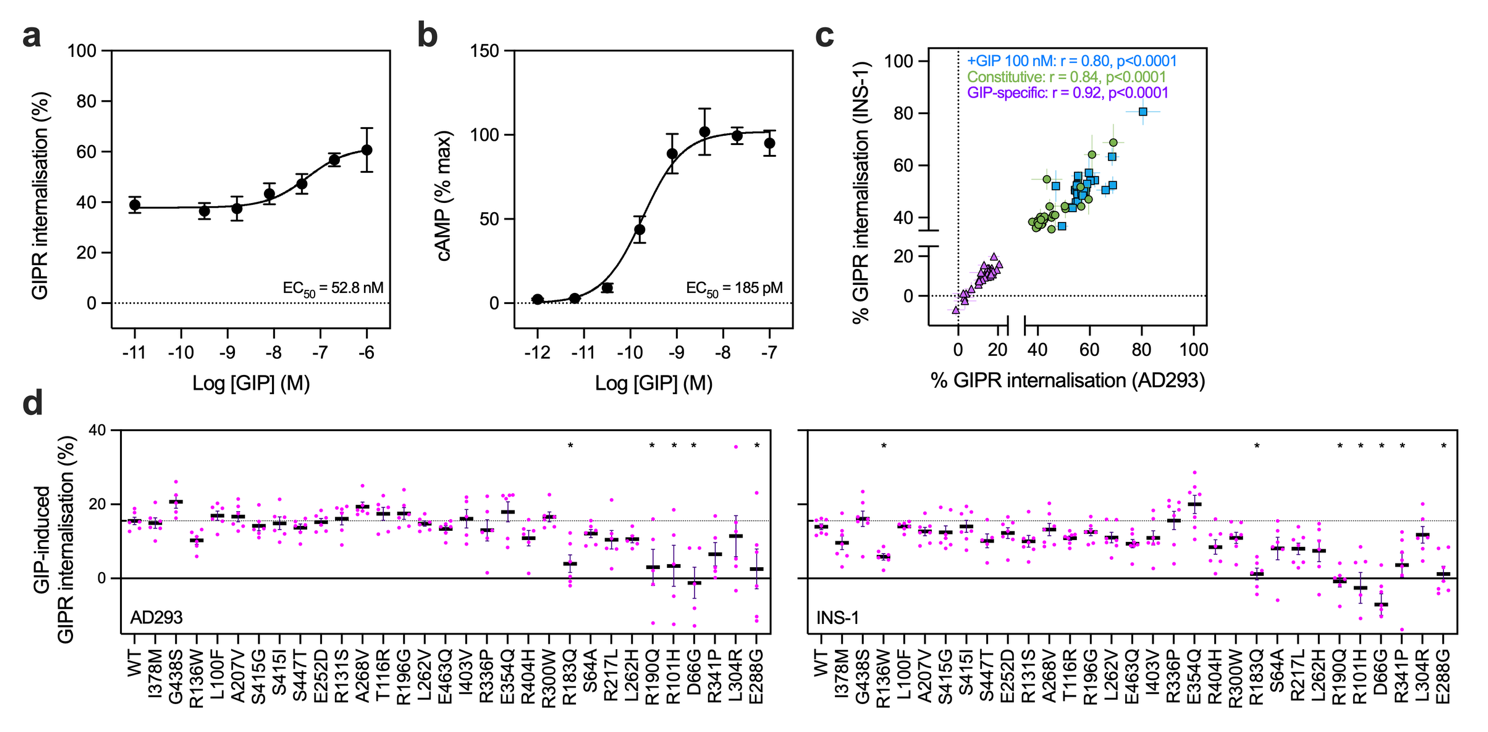

**Supplementary Figure 4.** (**a**) Wild-type SNAP-GIPR percentage internalisation concentration response, *n*=5. (**b**) Wild-type SNAP-GIPR cAMP concentration response, *n*=5. (**c**) Correlation between AD293 and INS-1 internalisation percentages. (**d**) Agonist-specific percentage GIPR internalisation in AD293 and INS-1 measured by subtracting vehicle from GIP response in Figure 4b and Figure 4c, with comparison by one-way matched ANOVA with Dunnett’s test *versus* wild-type. All data represented as mean ± SEM with individual data points shown in some cases.

**Supplementary Figure 5**

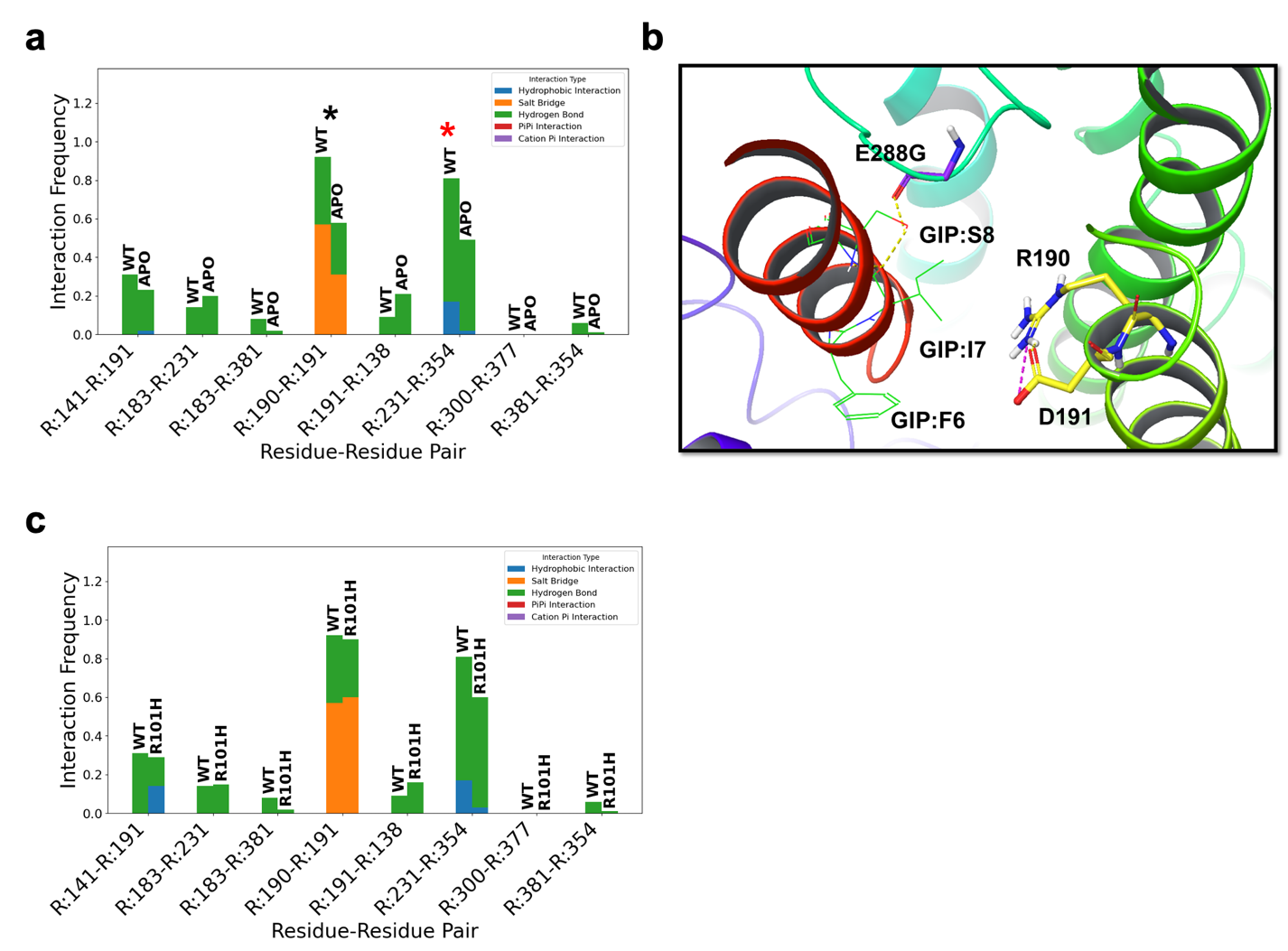

**Supplementary Figure 5.** (**a**) Wildtype receptor in the GIP bound and APO state. The intrahelical interaction in TM2 between neighbouring residues R190 and D191 is significantly enhanced in the presence of GIP. This suggests the setup of this local network, as well as TM4:Y231 TM6:E354 interhelical network is critical for GIP binding. (**b**) Representation of GIP (red ribbons) movement within the binding region in the presence of the E288G mutation. Interaction residues shown as stick representation. Hydrogen bonds are shown as yellow dashed lines and charge-charge interactions are shown as magenta dashed lines. (**c**) Mutation R101H show minor disruption of long-range protein-protein interactions at TM4-TM6. While the R101H mutation does not disrupt local orthosteric interactions typically observed in the presence of GIP, there is a small consistent long-range effect. These data together with other computational mutants suggest the TM6 E354 – TM4:Y231 inter-helical bundle interaction is very sensitive to changes in the receptor sequence. Since E354 is on TM6, directly implicated in signal transmission through the TM6 kink, even long distance (both sequentially and spatially) could affect G-protein engagement.

**Supplementary Figure 6**

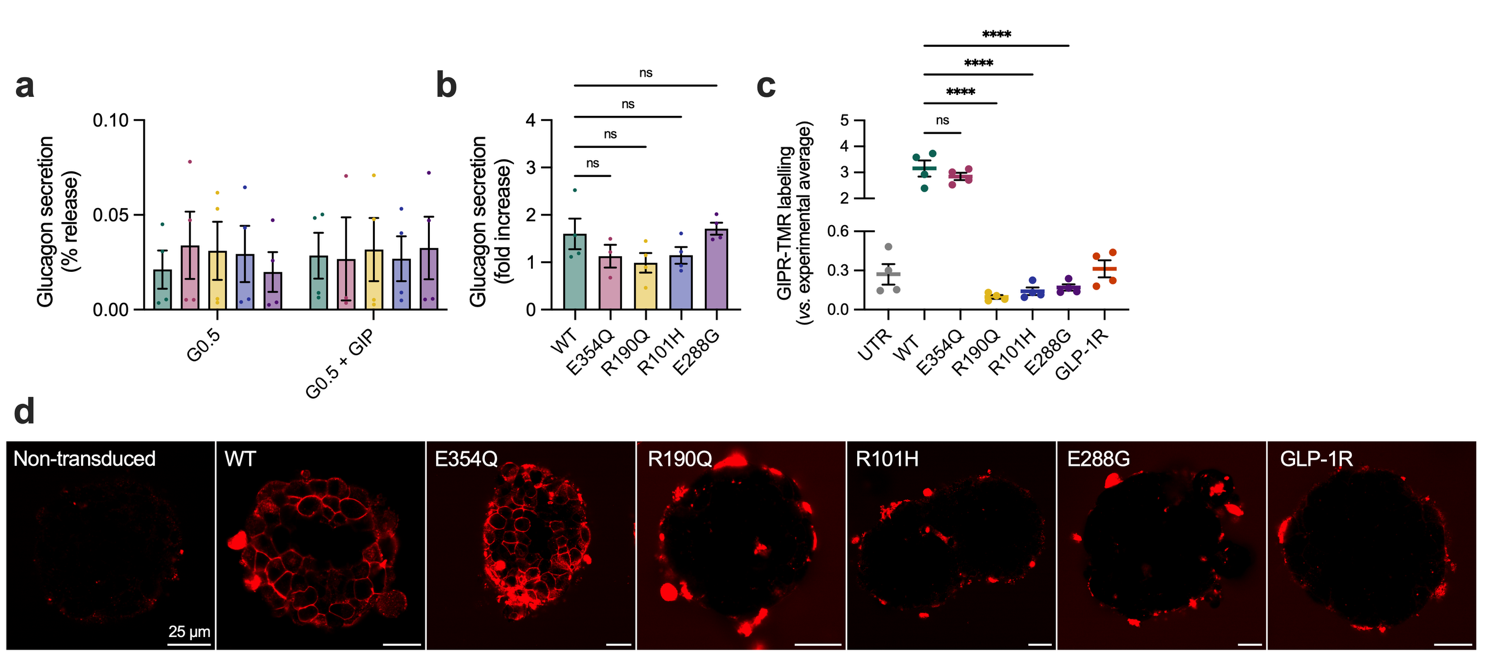

**Supplementary Figure 6.** (**a**) Release of glucagon from *Gipr*^-/-^ mouse islets transduced with GIPR adenoviruses at 0.5 mM glucose (G0.5), with and without 1,000 nM GIP, 30-minute stimulation, *n*=4. Wild-type and variant responses compared by two-way matched ANOVA with Dunnett’s test. (**b**) Data from (a) analysed as GIP-induced fold-change, with comparison using one-way matched ANOVA with Dunnett’s test. (**c**) Quantification of GIP-TMR labelling (1,000 nM, 30 minutes) in wild-type mouse islets transduced with SNAP-GIPR adenoviruses, *n*=4 repeats, comparison by one-way matched ANOVA with Dunnett’s test *versus* wild-type. (**d**) Representative images for data shown in (c); scale bars = 25 µm. *p<0.05, ****p<0.0001, using indicated statistical test. All data represented as mean ± SEM with individual data points.

**Supplementary Figure 7**

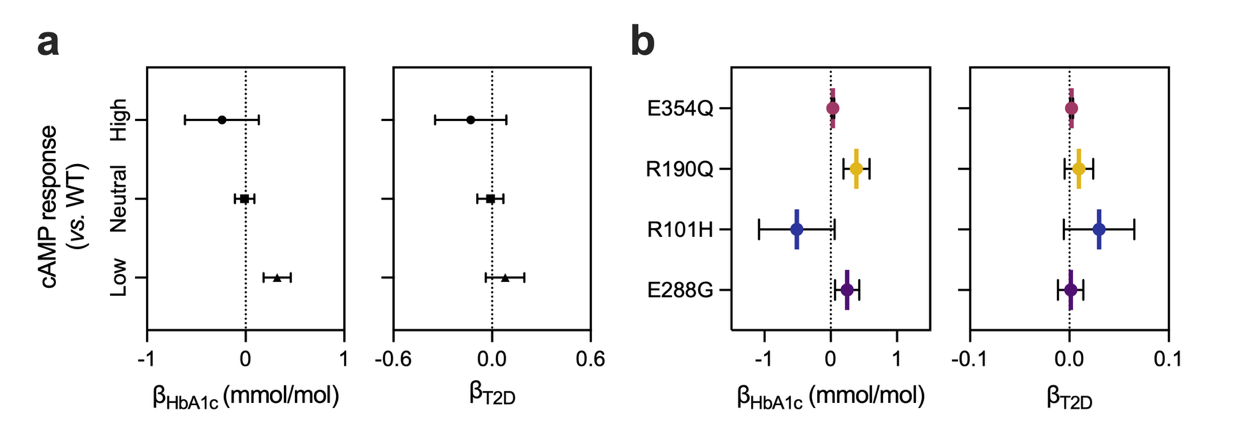

**Supplementary Figure 7.** (**a**) Results of burden test analysis, showing effects of *GIPR* variants grouped by cAMP response category on HbA1c and T2D risk, without adjustment for BMI. (**b**) Results of selected single variant association analyses, showing effects of E354Q, R190Q, R101H and E288G on HbA1c and T2D risk, without adjustment for BMI. Data are represented as mean ± 95% confidence intervals.
